# Plasma lipidomic profiles and risk of diabetes: two prospective cohorts of HIV-infected and HIV-uninfected individuals

**DOI:** 10.1101/2020.11.11.20229351

**Authors:** Eric Zhang, Jin Choul Chai, Amy A. Deik, Simin Hua, Anjali Sharma, Michael F. Schneider, Deborah Gustafson, David B. Hanna, Jordan E. Lake, Leah H. Rubin, Wendy S. Post, Kathryn Anastos, Todd Brown, Clary B. Clish, Robert C. Kaplan, Qibin Qi

## Abstract

**Objectives:** Antiretroviral therapy (ART) use is associated with disrupted lipid and glucose metabolism in people with HIV-infection. We aimed to identify plasma lipid species associated with risk of diabetes in the context of HIV infection.

**Research Design and Methods:** We profiled 211 plasma lipid species in 491 HIV-infected and 203 HIV-uninfected participants aged 35-55 years from the Women’s Interagency HIV study and the Multicenter AIDS Cohort Study. Cox proportional hazards model was used to examine associations between baseline lipid species and incident diabetes (166 diabetes cases were identified during a median follow-up of 12.6 years).

**Results:** We identified 11 lipid species, representing independent signals for 8 lipid classes/subclasses, associated with risk of diabetes (*P*<0.05 after FDR correction). After adjustment for multiple covariates, cholesteryl ester (CE)(22:4), lysophosphatidylcholine (LPC)(18:2), phosphatidylcholine (PC)(36:4), phosphatidylcholine-plasmalogen(34:3), and phosphatidylethanolamine (PE)(38:2) were associated with decreased risk of diabetes (HRs=0.70 to 0.82 per SD increment), while diacylglycerol(32:0), LPC(14:0), PC(38:3), PE(36:1), and triacylglycerol(50:1) were associated with increased risk of diabetes (HRs=1.26 to 1.56 per SD increment). HIV serostatus did not modify any lipid-diabetes associations; however, most of these lipid species were positively associated with HIV and/or ART use, including 3 diabetes-decreased (CE(22:4), LPC(18:2), PE(38:2)) and all 5 diabetes-increased lipid species.

**Conclusions:** This study identified multiple plasma lipid species associated with incident diabetes. Regardless of the directions of their associations with diabetes, most diabetes-associated lipid species were elevated in ART-treated people with HIV-infection. This suggests a complex role of lipids in the link between ART and diabetes in HIV infection.

## Introduction

Observed associations of antiretroviral therapy (ART) use, especially protease inhibitors (PI), with hyperglycemia, impaired glucose tolerance, and insulin resistance have raised concerns about diabetes in people with HIV infection (1-3). A higher incidence of diabetes has been reported in HIV-infected people compared to those without HIV infection in several (4,5), but not all (6) prospective HIV cohort studies. The pathogenesis of diabetes in HIV-infected people, which is not fully understood, may result from a complex interaction of chronic HIV infection, ART use, and factors unrelated to HIV such as diet, lifestyle, and genetic predisposition.(7,8)

Blood lipid disorders, such as high triglycerides and low high-density lipoprotein (HDL) cholesterol, are commonly present in both HIV infection (9) and diabetes (10). Earlier studies have focused on these conventional blood lipid measures which may ultimately encompass a large number of individual lipid species. Recent metabolomic and lipidomic studies in populations without HIV have identified multiple lipid species associated with diabetes risk, and suggested an important role of the structural features of acyl chains in the relationship between lipid species and diabetes (11-16). For example, short and saturated triacylglycerols, but not those with long and unsaturated acyl chains, were associated with increased diabetes risk (11,14,16). Our recent lipidomic profiling in two long-running HIV cohorts, the Women’s Interagency HIV Study (WIHS) and the Multicenter AIDS Cohort Study (MACS), identified multiple plasma lipid species elevated in HIV-infected people; and many of these elevated lipid species (e.g., ceramides, long and unsaturated triacylglycerols) were associated with increased risk of carotid artery atherosclerosis (17,18). Thus, lipidomics profiling may help us to better understand the biological link between HIV infection, disrupted lipid metabolism, and cardiometabolic disease. However to date, the relationship between plasma lipidomic profiles and diabetes risk has not been well-studied in the context of HIV-infection.

In the present study, we conducted a lipidomic analysis to examine associations between plasma lipidomic profiles including 211 lipid species and diabetes risk among 694 participants (491 HIV-infected, 203 HIV-uninfected) in the WIHS and MACS with a median follow-up of 12.6 years. We further examined the potential influences of HIV infection and related factors (e.g., ART use) on levels of lipid species and their associations with diabetes risk.

## Research Design and Methods

### Study Participants

Our current study is comprised of women from the WIHS and men from the MACS, two prospective multicenter cohort studies of individuals with or at high risk of HIV infection. The WIHS was initiated in 1994 and has enrolled over 4,000 women at six sites in the US. Recruitment in the WIHS occurred in three waves (1994-1995, 2001-2002, and 2010-2012) from HIV primary care clinics, hospital-based programs, community outreach, support groups, and other locations. The MACS was initiated in 1984, and has enrolled approximately 7,000 gay or bisexual men at four sites in the US. MACS recruitment also occurred in three waves (1984-1985, 1987, and 2002-2003). To ensure comparability with the HIV-positive participants, site recruitment targeted individuals who engaged in high-risk behaviors for enrollment in the HIV-negative group. WIHS and MACS participants undergo semiannual visits with a comprehensive physical examination, interviewer-administered questionnaire, and biological specimen collection at each visit. Additional details on the study design and methods have been described previously (19,20).

Beginning in 2004, the WIHS and MACS initiated a carotid artery imaging study scanning participants without prevalent CVD at 2-4 year intervals to ascertain progression of subclinical carotid artery atherosclerosis (21). In 2015, a metabolomics study was introduced to measure metabolomic profiles in plasma samples of participants collected during the baseline carotid artery imaging study (2004-2006) (22). Our current analysis includes 694 participants (391 women and 303 men), aged 35-55 years, who were free of CVD, carotid artery plaque, and diabetes at the baseline visit of the carotid artery imaging study. This metabolomics study was approved by the Institutional Review Board of Albert Einstein College of Medicine (Reference #: 009257). No identifiable personal data were used in this study. All necessary participant consent has been obtained and the appropriate institutional forms have been archived.

### Plasma lipidomic profiling

Lipidomic profiling was performed on stored frozen plasma specimens using liquid chromatography-tandem mass spectrometry (LC-MS) at the Broad Institute. Details on sample extraction, separation, and MS settings have been described elsewhere (23). Briefly, LC-MS data were acquired using a Nexera X2 U-HPLC (Shimadzu Corp.; Marlborough, MA) coupled to an Exactive Plus mass spectrometer (Thermo Fisher Scientific; Waltham, MA). Lipid identities were denoted by total acyl carbon number and total number of double bonds. Raw data were processed using TraceFinder software (Thermo Fisher Scientific; Waltham, MA) and Progenesis QI (Nonlinear Dynamics; Newcastle upon Tyne, UK). A total of 218 lipid species were quantified, and the median coefficient of variation (CV) was 4.7% (range, 2.8-31.4%), and 211 lipid species from 11 classes (16 classes/subclasses) were included in the analysis, after excluding 4 lipid species with CV ≥20%, and 3 lipid species with percentage of missing values ≥10%. Missing values due to below minimum detection level were imputed with ½ minimum values for a given lipid species. Detailed information on lipid classes/subclasses has been described elsewhere (18) and is also shown in **Supplementary Table 1**.

**Table 1.**
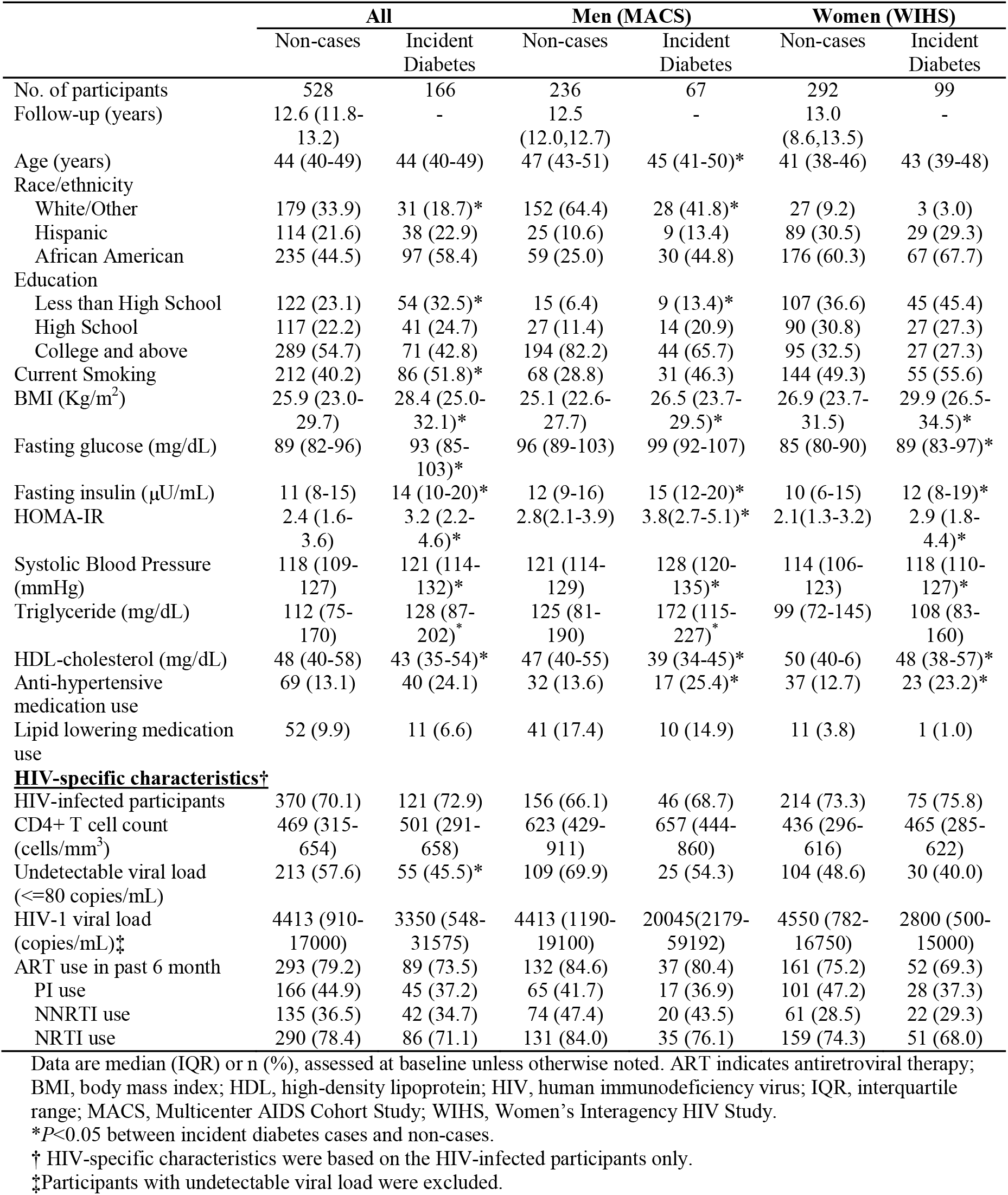
Participant characteristics at baseline

|  | **All** |  | **Men (MACS)** |  | **Women (WIHS)** |  |
| --- | --- | --- | --- | --- | --- | --- |
|  | Non-cases | Incident Diabetes | Non-cases | Incident Diabetes | Non-cases | Incident Diabetes |
| No. of participants | 528 | 166 | 236 | 67 | 292 | 99 |
| Follow-up (years) | 12.6 (11.8-13.2) | - | 12.5 (12.0,12.7) | - | 13.0 (8.6,13.5) | - |
| Age (years) | 44 (40-49) | 44 (40-49) | 47 (43-51) | 45 (41-50)* | 41 (38-46) | 43 (39-48) |
| Race/ethnicity |  |  |  |  |  |  |
| White/Other | 179 (33.9) | 31 (18.7)* | 152 (64.4) | 28 (41.8)* | 27 (9.2) | 3 (3.0) |
| Hispanic | 114 (21.6) | 38 (22.9) | 25 (10.6) | 9 (13.4) | 89 (30.5) | 29 (29.3) |
| African American | 235 (44.5) | 97 (58.4) | 59 (25.0) | 30 (44.8) | 176 (60.3) | 67 (67.7) |
| Education |  |  |  |  |  |  |
| Less than High School | 122 (23.1) | 54 (32.5)* | 15 (6.4) | 9 (13.4)* | 107 (36.6) | 45 (45.4) |
| High School | 117 (22.2) | 41 (24.7) | 27 (11.4) | 14 (20.9) | 90 (30.8) | 27 (27.3) |
| College and above | 289 (54.7) | 71 (42.8) | 194 (82.2) | 44 (65.7) | 95 (32.5) | 27 (27.3) |
| Current Smoking | 212 (40.2) | 86 (51.8)* | 68 (28.8) | 31 (46.3) | 144 (49.3) | 55 (55.6) |
| BMI (Kg/m<sup>2</sup>) | 25.9 (23.0-29.7) | 28.4 (25.0-32.1)* | 25.1 (22.6-27.7) | 26.5 (23.7-29.5)* | 26.9 (23.7-31.5) | 29.9 (26.5-34.5)* |
| Fasting glucose (mg/dL) | 89 (82-96) | 93 (85-103)* | 96 (89-103) | 99 (92-107) | 85 (80-90) | 89 (83-97)* |
| Fasting insulin (μU/mL) | 11 (8-15) | 14 (10-20)* | 12 (9-16) | 15 (12-20)* | 10 (6-15) | 12 (8-19)* |
| HOMA-IR | 2.4 (1.6-3.6) | 3.2 (2.2-4.6)* | 2.8(2.1-3.9) | 3.8(2.7-5.1)* | 2.1(1.3-3.2) | 2.9 (1.8-4.4)* |
| Systolic Blood Pressure (mmHg) | 118 (109-127) | 121 (114-132)* | 121 (114-129) | 128 (120-135)* | 114 (106-123) | 118 (110-127)* |
| Triglyceride (mg/dL) | 112 (75-170) | 128 (87-202)* | 125 (81-190) | 172 (115-227)* | 99 (72-145) | 108 (83-160) |
| HDL-cholesterol (mg/dL) | 48 (40-58) | 43 (35-54)* | 47 (40-55) | 39 (34-45)* | 50 (40-6) | 48 (38-57)* |
| Anti-hypertensive medication use | 69 (13.1) | 40 (24.1) | 32 (13.6) | 17 (25.4)* | 37 (12.7) | 23 (23.2)* |
| Lipid lowering medication use | 52 (9.9) | 11 (6.6) | 41 (17.4) | 10 (14.9) | 11 (3.8) | 1 (1.0) |
| **HIV-specific characteristics†** |  |  |  |  |  |  |
| HIV-infected participants | 370 (70.1) | 121 (72.9) | 156 (66.1) | 46 (68.7) | 214 (73.3) | 75 (75.8) |
| CD4+ T cell count (cells/mm<sup>3</sup>) | 469 (315-654) | 501 (291-658) | 623 (429-911) | 657 (444-860) | 436 (296-616) | 465 (285-622) |
| Undetectable viral load (<=80 copies/mL) | 213 (57.6) | 55 (45.5)* | 109 (69.9) | 25 (54.3) | 104 (48.6) | 30 (40.0) |
| HIV-1 viral load (copies/mL)‡ | 4413 (910-17000) | 3350 (548-31575) | 4413 (1190-19100) | 20045(2179-59192) | 4550 (782-16750) | 2800 (500-15000) |
| ART use in past 6 month | 293 (79.2) | 89 (73.5) | 132 (84.6) | 37 (80.4) | 161 (75.2) | 52 (69.3) |
| PI use | 166 (44.9) | 45 (37.2) | 65 (41.7) | 17 (36.9) | 101 (47.2) | 28 (37.3) |
| NNRTI use | 135 (36.5) | 42 (34.7) | 74 (47.4) | 20 (43.5) | 61 (28.5) | 22 (29.3) |
| NRTI use | 290 (78.4) | 86 (71.1) | 131 (84.0) | 35 (76.1) | 159 (74.3) | 51 (68.0) |

Data are median (IQR) or n (%), assessed at baseline unless otherwise noted. ART indicates antiretroviral therapy; BMI, body mass index; HDL, high-density lipoprotein; HIV, human immunodeficiency virus; IQR, interquartile range; MACS, Multicenter AIDS Cohort Study; WIHS, Women's Interagency HIV Study.
\* $P < 0.05$ between incident diabetes cases and non-cases.
† HIV-specific characteristics were based on the HIV-infected participants only.
‡ Participants with undetectable viral load were excluded.

### Assessments of HIV infection, diabetes, and other variables

Data on demographic, behavioral, clinical, and laboratory variables were collected using standardized protocols at semi-annual visits (21). HIV infection was ascertained by enzyme-linked immunosorbent assay (ELISA) and confirmed by Western blot. HIV-specific parameters including CD4+ T-cell counts, HIV-1 viral load, and detailed information on classes of ART drugs (protease inhibitors [PI], non-nucleoside reverse transcriptase inhibitors [NNRTI] and nucleoside reverse transcriptase inhibitors [NRTI] have been described previously (24).

Incident diabetes was defined as the first time a study participant was observed with fasting glucose ≥126 mg/dL, HbA1C ≥6.5%, or self-report of anti-diabetic medication during follow up. Fasting glucose, fasting insulin and HbA1c were measured using standard assays at central laboratories of the WIHS (Quest Diagnostics, Baltimore, MD) (5,25) and MACS (Heinz Laboratory, Pittsburgh, PA) (26,27). Insulin resistance was estimated using the homeostasis model assessment (HOMA-IR) equation, defined as (insulin [μU/mL] × glucose [mg/dL])/405 (28). Measurements of other cardiometabolic traits including body mass index (BMI), systolic blood pressure, high-density lipoprotein (HDL)-cholesterols and triglycerides have been described previously (29).

### Statistical Analysis

Baseline characteristics of those with incident diabetes were compared to those without incident diabetes using the Mann–Whitney *U* test (continuous variables) and χ^2^ test (categorical variables) overall, and separately in men and women. Sex-specific inverse normal transformations were performed on each lipid species before analyses.

Associations of plasma lipidomic profiles with risk of diabetes were examined in the following steps. First, multivariate Cox proportional hazards models (referred to as Cox model afterwards) stratified by sex were used to examine the association between each of the 211 lipid species and risk of diabetes, adjusting for age, race/ethnicity, education and current smoking status (basic model). False discovery rate (FDR) was applied to control for multiple testing (30). Second, a lipid class/subclass specific analysis was conducted. This focused on lipid class/subclasses for which at least one lipid species in this class/subclass was significantly associated with risk of diabetes after FDR correction. Lipid species which showed the strongest association (smallest *P*-value) with risk of diabetes within each lipid class/subclass was selected as a representative lipid species (top lipid species) of correlated lipid species in the same class/subclass. If there were additional statistically significant lipid species not highly correlated with the top lipid (Pearson correlation<0.8) within the same class/subclass, we further conducted a joint analysis which included the top and each of the other lipids in the Cox model simultaneously. This lipid class/subclass specific analysis allowed us to identify additional signals represented by secondary lipid species associated with risk of diabetes. Finally, for the featured lipid species (both top and secondary lipid species), the multivariate Cox model was further adjusted for HIV serostatus and ART use (HIV-uninfected, HIV-infected on ART, HIV-infected ART-naive), CD4+ T cell counts, and cardiometabolic risk factors including BMI, systolic blood pressure, HDL cholesterols, triglycerides, and use of antihypertensive and lipid-lowering medications. Analyses were also conducted separately within HIV-infected and HIV-uninfected participants as well as within men and women. Possible effect modifications of lipid species by HIV infection or by sex were examined by including an interaction term in the Cox model. In addition, we also combined results from men and women by fixed-effect meta-analysis.

Partial Spearman correlation analysis was used to examine correlation coefficients of lipids species with fasting glucose, fasting insulin, HOMA-IR, and other cardiometabolic traits, and HIV parameters (CD4+ T cell counts and viral load), after adjusting for age and sex. Linear regression was used to compare lipid species levels between HIV-infected and HIV-uninfected participants; and across three groups according to HIV status and current ART use (HIV-uninfected, HIV-infected on ART, HIV-infected ART-naive), adjusting for age and sex. In addition, the associations between lipid species and three different classes of ART medications, including PI, NNRTI, and NRTI use, were also examined. All analyses were conducted in R 3.4.0 (*The R Foundation for Statistical Computing, 2017*).

## Results

### Participant Characteristics

In 694 participants (303 men and 391 women), we identified 166 incident diabetes cases (67 men and 99 women) during a median follow-up of 12.6 (IQR: 11.8, 13.2) years. **Table 1** shows baseline characteristics of these participants. As compared to women, men were older, better educated, and more likely to be white and have lower BMI, and were less likely to smoke. The contrast between incident diabetes cases and non-cases appeared to share a similar pattern for men and women: that is, participants who developed diabetes were more likely to be African Americans and smokers, were less educated, had higher levels of BMI, systolic blood pressure and triglycerides, and lower levels of HDL cholesterols. Baseline demographic, socioeconomic and behavioral variables between the HIV-infected (n=491) and HIV-uninfected (n=203) groups were generally similar and comparable (**Supplementary Table 2**), which is in line with the design of WIHS and MACS (19,20).

**Table 2.** Associations of 11 featured lipid species with risk of diabetes

| Lipid species | All (N=694) |  | HIV+ (N=491) |  | HIV- (N=203) |  | P for interaction |
| --- | --- | --- | --- | --- | --- | --- | --- |
|  | HR (95%CI) | P-value | HR (95%CI) | P-value | HR (95%CI) | P-value |  |
| **CE (22:4)** |  |  |  |  |  |  |  |
| Model 1 | 0.63 (0.54,0.75) | <0.001 | 0.65 (0.54,0.78) | <0.001 | 0.54 (0.37,0.79) | <0.001 | 0.69 |
| Model 2 | 0.70 (0.57,0.85) | <0.001 | 0.68 (0.54,0.84) | <0.001 | 0.85 (0.49,1.46) | 0.55 | 0.59 |
| **DAG (32:0)** |  |  |  |  |  |  |  |
| Model 1 | 1.61 (1.37,1.90) | <0.001 | 1.61 (1.34,1.95) | <0.001 | 1.59 (1.15,2.20) | <0.001 | 0.73 |
| Model 2 | 1.56 (1.27,1.93) | <0.001 | 1.65 (1.29,2.10) | <0.001 | 1.16 (0.75,1.80) | 0.49 | 0.38 |
| **LPC (18:2)\*** |  |  |  |  |  |  |  |
| Model 1 | 0.72 (0.60,0.86) | <0.001 | 0.76 (0.62,0.93) | <0.001 | 0.62 (0.43,0.88) | <0.001 | 0.44 |
| Model 2 | 0.79 (0.65,0.96) | 0.02 | 0.84 (0.67,1.05) | 0.14 | 0.74 (0.47,1.16) | 0.19 | 0.54 |
| **LPC (14:0)\*** |  |  |  |  |  |  |  |
| Model 1 | 1.41 (1.18,1.69) | <0.001 | 1.30 (1.04,1.62) | 0.01 | 1.71 (1.21,2.41) | <0.001 | 0.31 |
| Model 2 | 1.28 (1.06,1.55) | 0.01 | 1.20 (0.96,1.51) | 0.1 | 1.35 (0.93,1.96) | 0.11 | 0.5 |
| **PC (P-34:3)** |  |  |  |  |  |  |  |
| Model 1 | 0.74 (0.63,0.87) | <0.001 | 0.74 (0.61,0.90) | <0.001 | 0.74 (0.54,1.00) | 0.05 | 0.81 |
| Model 2 | 0.79 (0.65,0.96) | 0.02 | 0.76 (0.60,0.97) | 0.02 | 0.89 (0.60,1.34) | 0.59 | 0.59 |
| **PC (38:3)\*** |  |  |  |  |  |  |  |
| Model 1 | 1.44 (1.19,1.73) | <0.001 | 1.30 (1.05,1.60) | 0.01 | 2.00 (1.34,2.98) | <0.001 | 0.12 |
| Model 2 | 1.26 (1.02,1.56) | 0.02 | 1.15 (0.90,1.48) | 0.23 | 1.60 (1.00,2.56) | 0.04 | 0.15 |
| **PC (36:4)\*** |  |  |  |  |  |  |  |
| Model 1 | 0.76 (0.64,0.90) | <0.001 | 0.85 (0.70,1.03) | 0.11 | 0.56 (0.40,0.78) | <0.001 | 0.04 |
| Model 2 | 0.82 (0.68,0.98) | 0.03 | 0.90 (0.72,1.13) | 0.39 | 0.60 (0.39,0.91) | 0.01 | 0.09 |
| **PE (36:1)\*** |  |  |  |  |  |  |  |
| Model 1 | 1.46 (1.22,1.76) | <0.001 | 1.48 (1.18,1.84) | <0.001 | 1.42 (1.03,1.96) | 0.03 | 0.83 |
| Model 2 | 1.37 (1.11,1.69) | <0.001 | 1.42 (1.09,1.86) | <0.001 | 1.17 (0.78,1.74) | 0.43 | 0.95 |
| **PE (38:2)\*** |  |  |  |  |  |  |  |
| Model 1 | 0.70 (0.58,0.83) | <0.001 | 0.74 (0.60,0.91) | <0.001 | 0.61 (0.44,0.86) | <0.001 | 0.27 |
| Model 2 | 0.76 (0.63,0.93) | <0.001 | 0.83 (0.66,1.04) | 0.12 | 0.66 (0.46,0.95) | 0.02 | 0.23 |
| **PE (P-38:3)** |  |  |  |  |  |  |  |
| Model 1 | 0.79 (0.67,0.93) | <0.001 | 0.80 (0.66,0.97) | 0.02 | 0.76 (0.54,1.07) | 0.12 | 0.92 |
| Model 2 | 0.89 (0.74,1.06) | 0.21 | 0.89 (0.72,1.10) | 0.30 | 0.92 (0.64,1.34) | 0.69 | 0.91 |
| **TAG (50:1)** |  |  |  |  |  |  |  |
| Model 1 | 1.56 (1.32,1.83) | <0.001 | 1.57 (1.30,1.89) | <0.001 | 1.51 (1.08,2.12) | 0.01 | 0.65 |
| Model 2 | 1.48 (1.20,1.83) | <0.001 | 1.59 (1.25,2.02) | <0.001 | 0.94 (0.58,1.53) | 0.82 | 0.28 |

Data are Hazard ratios (HR) and 95% confidence intervals (CI) of incident diabetes per SD increment of lipid species levels, stratified by sex and adjusted for age, race/ethnicity, education, study site, current smoking, HIV serostatus and treatment status (HIV-uninfected, HIV-infected ART using, HIV- infected ART naive) and CD4 cell counts (Model 1); and further adjusted for systolic blood pressure, HDL-cholesterol, triglycerides, BMI, anti-hypertensive medication use and lipid lowering medication use (Model 2).
\*Top and Secondary lipid species within the same lipid classes/subclasses were included in the Cox models simultaneously to estimate HRs (95% CI) and *P*-values
CE, cholesteryl ester; DAG, diacylglycerol; LPC, lysophosphatidylcholine; PC, phosphatidylcholine; PC(P), phosphatidylcholine plasmalogen; PE, phosphatidylethanolamine; PE (P), phosphatidylethanolamine plasmalogen; TAG, triacylglycerol.

### Lipidomic profiles and incident diabetes

Among 211 individual lipid species examined in this analysis, 80 lipid species from 6 lipid classes (8 classes/subclasses), including cholesteryl esters (CEs), diacylglycerols (DAGs), lysophosphatidylcholines (LPCs), phosphatidylcholines (PCs) (including PC and PC plasmalogen [PC-P] subclasses), phosphatidylethanolamines (PEs) (including PE and PE plasmalogen [PE-P] subclasses), and triacylglycerol (TAGs), showed significant associations with risk of diabetes after FDR adjustment (**Supplementary Figure 1 and Supplementary Table 3**). The top lipid species were CE (22:4), DAG (32:0), LPC (18:2), PC (P-34:3), PC (38:3), PE (36:1), PE (P-38:3) and TAG (50:1). We further plotted the association results of all lipid species in these 8 classes/subclasses with risk of diabetes by numbers of carbons and double-bonds of the acyl chains **(Figure 1)**. Almost all the CEs, PCs-P, and PEs-P showed inverse associations with risk of diabetes, while all DAGs and TAGs showed positive associations with risk of diabetes. DAGs with short and saturated or mono-unsaturated (0-1 double bonds) acyl chains and TAGs with medium and saturated or mono-unsaturated acyl chains showed the strongest associations with risk of diabetes. In addition to individual lipid species analyses, we also examined associations of lipid classes/subclasses with risk of diabetes. DAG and TAG lipid classes, especially those with saturated or mon-unsaturated acyl chains, were significantly associated with risk of diabetes (**Supplementary Table 4**).

**Figure 1.**
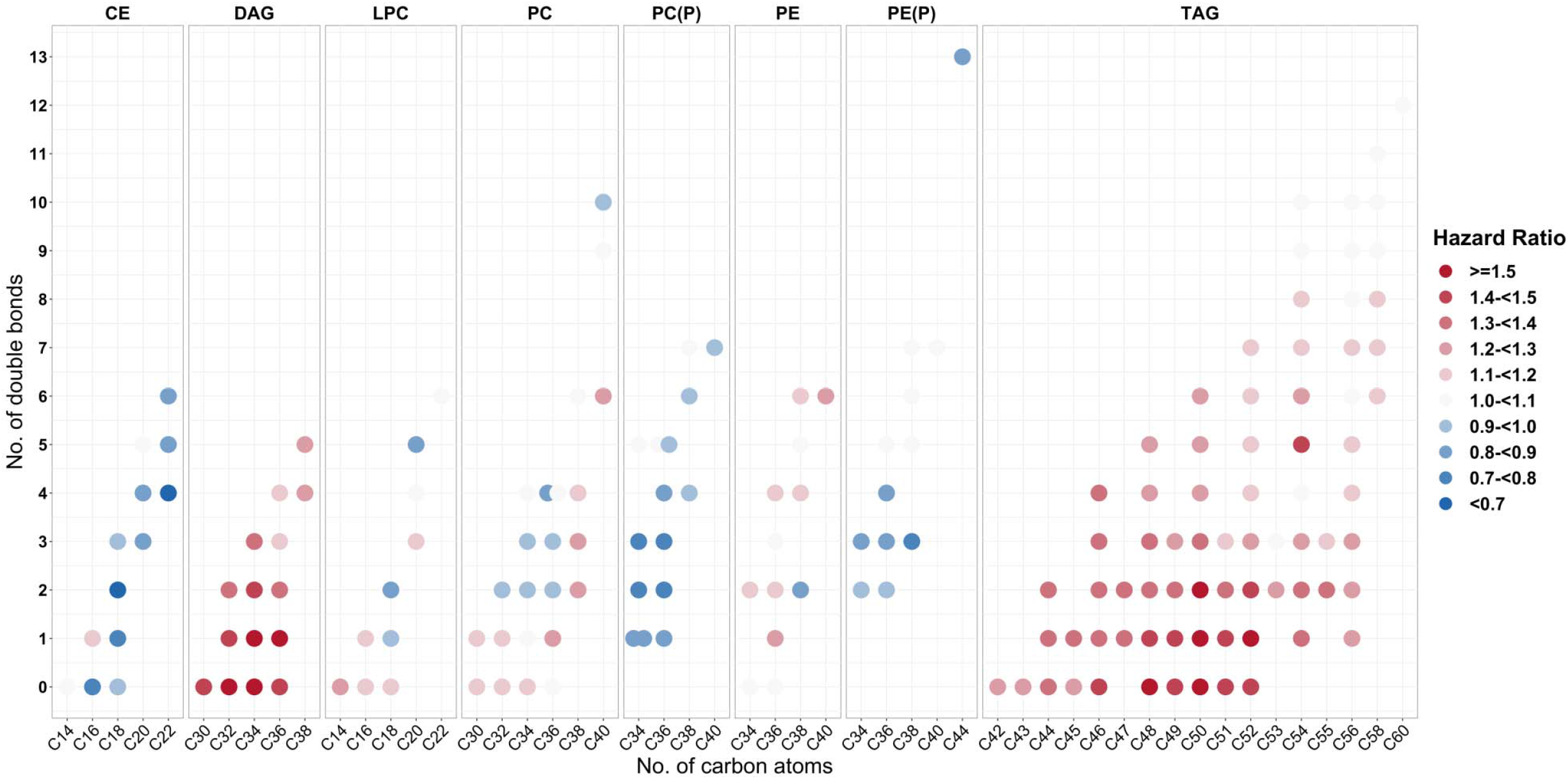
Associations of individual lipid species from 8 lipid classes/subclasses with risk of diabetes. Individual lipid species are depicted by filled circles and arranged by lipid classes/subclasses according to the number of total carbon atoms (x-axis) and number of double bonds (y-axis). Data are hazard ratios for risk of diabetes per 1-SD increment of lipid species levels, stratified by sex and adjusted for age, race/ethnicity, education, study site and current smoking. CE, cholesteryl ester; DAG, diacylglycerol; LPC, lysophosphatidylcholine; PC, phosphatidylcholine; PC(P), phosphatidylcholine plasmalogen; PE, phosphatidylethanolamine; PE (P), phosphatidylethanolamine plasmalogen; TAG, triacylglycerol.

Through the joint analyses, we identified 3 secondary lipid species, LPC (14:0), PC (36:4), and PE (38:2), associated with risk of diabetes, independent of the top lipid species in the respective lipid classes/subclasses (**Supplementary Figure 2**). All 3 secondary lipid species had opposite associations with risk of diabetes compared to the top lipid species in the same classes/subclasses, although a moderate level of positive correlations (r=0.2 to 0.4) were observed between the top and secondary lipid species (**Supplementary Table 5**). For example, the top lipid species LPC(18:2) was inversely associated with risk of diabetes (HR=0.72) while the secondary lipid LPC(14:0) had a positive association with risk of diabetes (HR=1.40).

**Figure 2.**
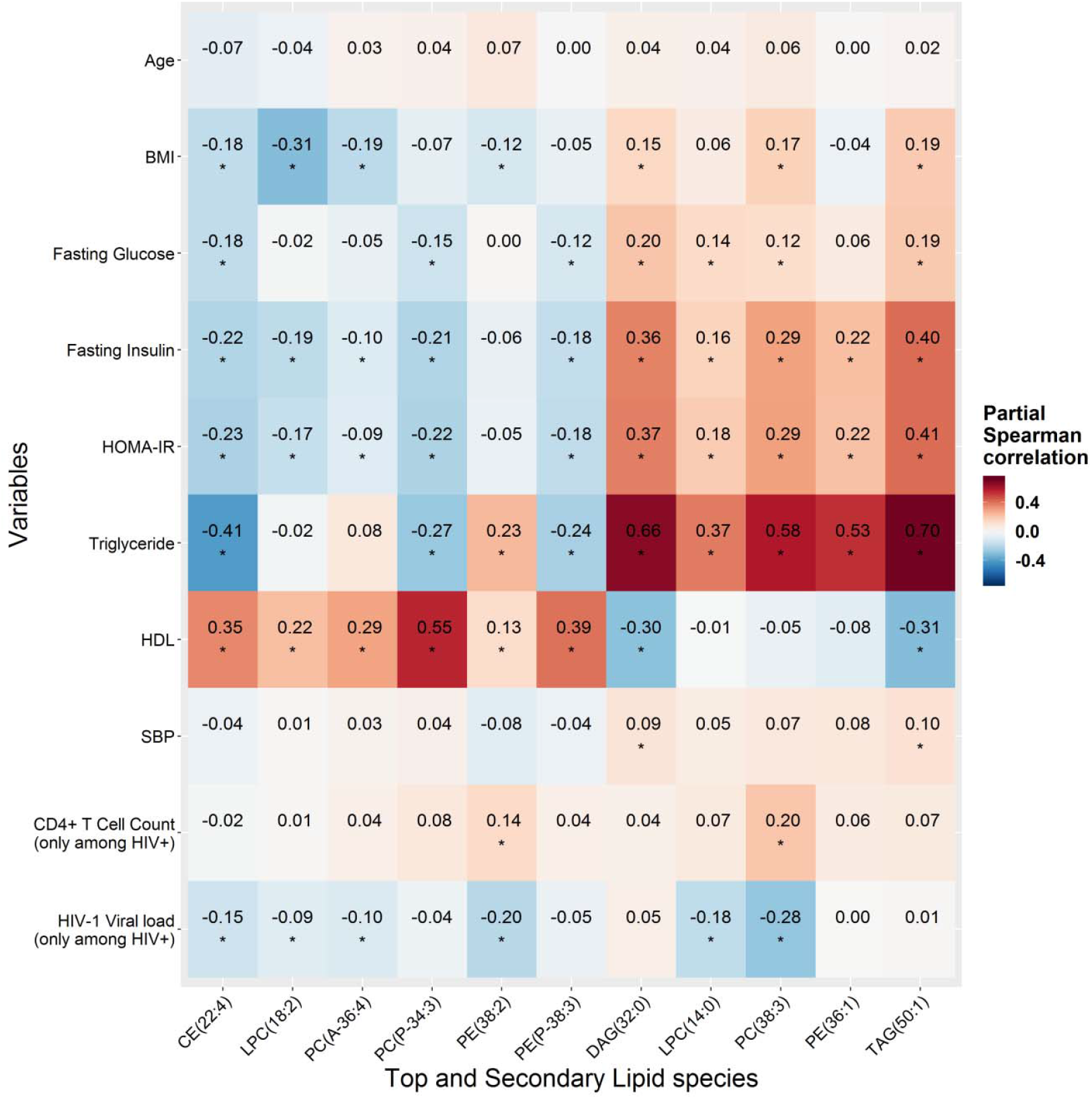
Partial Spearman correlation of lipid species with diabetes and HIV related factors. Data are partial Spearman correlation coefficients (r) of 11 diabetes-associated lipid species levels with age, BMI, fasting glucose, fasting insulin, HOMR-IR, HDL-cholesterol and triglycerides (excluding participants taking lipid-lowering medication); systolic blood pressure (excluding participants taking anti-hypertensive medication); and baseline CD4+ T cell count and HIV-1 viral load (among people with HIV infection only), adjusting for age and sex. \**P*-value <0.05. BMI, body mass index; CE, cholesteryl ester; DAG, diacylglycerol; LPC, lysophosphatidylcholine; PC, phosphatidylcholine; PC(P), phosphatidylcholine plasmalogen; PE, phosphatidylethanolamine; PE (P), phosphatidylethanolamine plasmalogen; TAG, triacylglycerol.

The multivariate adjusted hazards ratios (HRs) of the 11 featured (8 top and 3 secondary) lipid species with risk of diabetes are shown in **Table 2**. Six lipid species, CE (22:4), LPC (18:2), PC (P-34:3), PC (36:4), PE (38:2) and PE (P-38:3) were associated with decreased risk of diabetes (HRs=0.63 to 0.79 per SD increment), while 5 lipid species, DAG (32:0), LPC (14:0), PC (38:3), PE (36:1) and TAG (50:1) were associated with increased risk of diabetes (HRs=1.41 to 1.61), after adjustment for demographic, socioeconomic, behavioral, and HIV infection and related factors. After additional adjustment for traditional cardiometabolic risk factors, the strength of the associations between lipid species and risk of diabetes was somewhat attenuated, but all except PE (P-38:3) remained statistically significant.

There was little evidence for effect modification by HIV on associations of these 11 lipid species and risk of diabetes. Only PC (36:4) showed a stronger association with risk of diabetes in HIV-uninfected individuals compared to those with HIV infection (raw *P* for interaction=0.04) (**Table 2**). Associations between these lipid species and risk of diabetes were generally consistent between men and women, except for PE (P-38:3) which showed a stronger association with risk of diabetes in men compared to women (raw *P* for interaction=0.02) (**Supplementary Table 6**). However, none of these interaction terms remained significant after FDR adjustment. Similar results were obtained when using a meta-analysis approach to combine results from men and women and test potential sex-differences in associations between lipid species and risk of diabetes (**Supplementary Table 6)**.

Considering potential inaccuracy of using HbA1c test among HIV-infected people,(26) we performed a sensitivity analysis by removing HbA1c criteria from the definition of diabetes. This new definition led to a reduction of 14 incident diabetes cases, but the overall patterns of associations between lipid species and risk of diabetes remained the same (data not shown).

### Diabetes-associated lipid species and cardiometabolic risk factors

**Figure 2** shows correlations between 11 diabetes-associated lipid species and traditional cardiometabolic risk factors. The 6 lipid species that were associated with decreased risk of diabetes generally showed favorable correlations with adiposity (low BMI), glycemic traits (low levels of fasting glucose, fasting insulin, and HOMA-IR), and blood lipids (low triglycerides, high HDL-cholesterol), though there was a weak-to-moderate positive correlation between PE (P-38-3) and triglycerides. The 5 lipid species which were associated with increased risk of diabetes, showed unfavorable correlations with these traditional cardiometabolic risk factors, with strongest correlations observed for high triglycerides.

Correlation patterns between these lipid species and cardiometabolic risk factors were generally similar between HIV-infected and HIV-uninfected individuals, except for PC (36:4) and PE (38:2) which showed significant inverse correlations with fasting insulin and HOMA-IR only in HIV-uninfected but not in HIV-infected individuals **(Supplementary Figure 3)**. Among HIV-infected individuals, a few weak-to-moderate correlations were observed between some of these diabetes-associated lipid species and CD4 T cell counts and/or HIV-1 viral load (**Figure 2)**.

**Figure 3.**
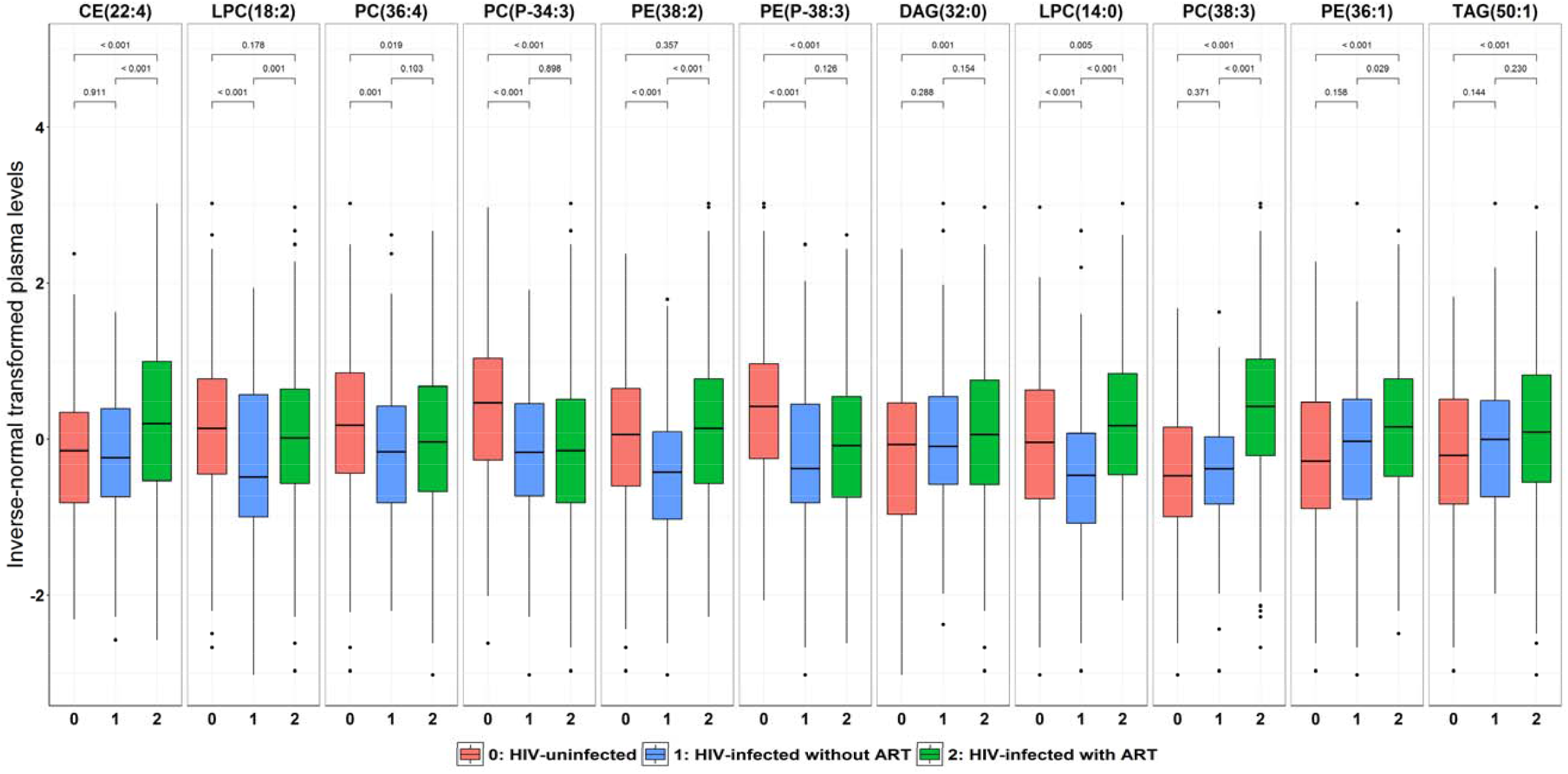
Plasma levels of diabetes-associated lipid species according to HIV infection and ART use. Data are inverse normal transformed levels of 11 diabetes-associated lipid species in HIV-uninfected individuals (red), HIV-infected ART-naïve (blue), and HIV-infected using ART (green). P-values for comparisons between groups were calculated using Wilcoxon rank sum test. ART indicates antiretroviral therapy; CE, cholesteryl ester; DAG, diacylglycerol; LPC, lysophosphatidylcholine; PC, phosphatidylcholine; PC(P), phosphatidylcholine plasmalogen; PE, phosphatidylethanolamine; PE (P), phosphatidylethanolamine plasmalogen; TAG, triacylglycerol.

### HIV infection, ART use and diabetes-associated lipid species

We then examined the relationships of these diabetes-associated lipid species with HIV infection and ART use (**Figure 3** and **Supplementary Table 7**). Among 6 lipid species associated with decreased risk of diabetes, PC (36:4), PC (P-34:3) and PE (P-38:3) were significantly lower in HIV-infected individuals (using or not using ART combined) compared to HIV-uninfected individuals, and there were no significant differences between HIV-infected individuals using or not using ART. Plasma LPC (18:2) was significantly lower in HIV-infected compared to HIV-uninfected individuals, but the levels were elevated in HIV-infected individuals using ART compared to those not using ART. Plasma CE (22:4) was significantly higher in HIV-infected compared to HIV-uninfected individuals, which was primarily driven by elevated levels in HIV-infected individuals using ART. PE(38:2) showed no significant differences between HIV-infected and HIV-uninfected individuals, but the levels were significantly higher in HIV-infected using ART compared to those not using ART.

Among the five lipid species associated with increased risk of diabetes, DAG(32:0), PC(38:3), PE(36:1), and TAG(50:1) were significantly higher in HIV-infected compared to HIV-infected individuals, with the highest levels in HIV-infected individuals on ART (**Figure 3**). Although the overall comparison between HIV-infected and HIV-uninfected individuals did not show significant difference in LPC(14:0), the levels were significantly higher in HIV-infected individuals using ART compared to HIV-infected individuals not using ART and HIV-uninfected individuals.

Further analyses by different classes of ART drugs showed that PI-based ART had the most significant influences on plasma levels of these lipid species. PI use was associated with elevated levels of 8 lipid species, NRTI use was associated with elevated levels of 6 lipid species, and NNRTI use was associated with elevated levels of 3 lipid species (**Supplemental Figure 4**).

## Discussion

In the combined two cohorts of 694 men and women including HIV-infected and HIV-uninfected individuals, 80 out of 211 lipid species examined from 16 lipid classes were identified to be individually associated with risk of diabetes. Because most lipid species in the same lipid class/subclass were correlated, a lipid-class/subclass specific analysis was conducted to further examine the independent effects of lipid species on risk of diabetes, revealing 11 featured lipid species (8 top and 3 secondary) from 8 lipid classes/subclasses. The observed associations between these lipid species and diabetes risk were supported by their correlations with traditional risk factors. However, their associations with risk of diabetes, except for PE (P-38:3), were only slightly attenuated after controlling for traditional risk factors, suggesting potentially independent effects of these lipid species on risk of diabetes.

Consistent with previous studies in non-HIV cohorts (11-16), our individual lipid species analyses indicated that almost all the CEs, PC(P)s and PE(P)s were inversely associated with risk of diabetes, while many PCs and PEs, and all DAGs and TAGs were positively associated with risk of diabetes. Notably, after controlling for the top lipid species within each class/subclass, we identified three secondary lipid species (LPC [14:0], PC [36:4] and PE [38:02]) which showed opposite associations with risk of diabetes, compared to the top lipid species (LPC[18:2], PC[38:3], and PE [36:1]. Before adjusting for the top lipid species, associations of these secondary lipid species with risk of diabetes could have been masked because of their positive correlations with the top lipid species. Consistent with this idea, two previous studies in non-HIV cohorts also found all TAGs to be associated with a higher risk of diabetes, while after controlling for total TAG content, several TAGs showed inverse associations with risk of diabetes (14,16). We found similar results for the TAG-specific analyses, although the emerging inverse associations did not reach significance. These data suggested that correlations among lipid species need to be taken into account in the analyses.

In line with our results on PC(P)s and PE(P)s, plasmalogens have been suggested to have antioxidical, antiapoptotic and anti-inflammatory functions which may lower risk of diabetes (31,32). LPCs may improve glucose metabolism through enhanced insulin sensitivity and anti-inflammation (33-35). Alteration in skeletal muscle phospholipid metabolism (increased PCs and decreased PEs) due to *CEPT1* deficiency has been related to improved insulin sensitivity in high-fat diet–fed mice (36). However, the biological mechanisms underlying the associations between individual lipid species and risk of diabetes remains unclear. Our data suggest that lipid species within the same class/subclass, even though they are correlated with each other, may have different biological links with diabetes due to structural features of the acyl chains.

This study included both HIV-infected and HIV-uninfected individuals, but we did not find strong evidence for HIV-serostatus modification on the associations between lipid species and risk of diabetes. Only one lipid species PC(36:4) showed a suggestive interaction with HIV infection (although it did not pass FDR correction), with a significant inverse association with risk of diabetes in HIV-uninfected individuals but not in HIV-infected. Consistently, PC(36:4) showed inverse correlations with insulin resistance markers only in HIV-uninfected but not in HIV-infected individuals. Previous studies in non-HIV cohorts also showed the inverse associations of PC(36:4) with insulin resistance (37) and type 2 diabetes (15). We observed a significantly lower level of PC(36:4) in HIV-infected compared to HIV-uninfected individuals, but it is unknown whether this might influence its association with risk of diabetes in HIV-infected people.

A unique contribution of this study is that we investigated the relationships of HIV infection and ART use with diabetes-associated lipid species and found some intriguing results. For the five lipid species associated with increased risk of diabetes, their plasma levels were all elevated in HIV-infected individuals on ART, especially in those with PI use. This is consistent with reports that PI-based ART may have more detrimental effects on lipid metabolism and glucose metabolism compared with other ART regimens (1-3,7,8). The observed relationships of these lipid species with ART use and risk of diabetes are in the expected directions (i.e., elevated lipid species in ART use were associated with higher risk of diabetes), which supports one of the pathophysiological hypotheses that ART use may disrupt lipid metabolism, and thus contribute to insulin resistance and increased risk of diabetes (7,8). However, interestingly, we found that several lipid species (i.e., CE[22:4], LPC[18:2] and PE[38:2]) which were associated with lower risk of diabetes, were also elevated in HIV-infected individuals using ART. Although further validation is needed, these unexpected results indicates that ART use might increase levels of some lipid species which could be beneficial for diabetes, suggesting a complex role of ART use in the development of diabetes among HIV-infected individuals (8).

To the best of our knowledge, this study is the first prospective study of lipidomics and risk of diabetes in the context of HIV infection. Major strengths of the study include well-characterized HIV-cohorts with both HIV-infected and HIV-uninfected participants, high-throughput and high-quality lipidomic profiling, a long-term follow-up on diabetes status and multiple cardiometabolic measures which may provide some information on plausible mechanisms. However, this study has several limitations. Due to the nature of observational study design, this study cannot make causal inferences. Our untargeted lipidomics had a broad coverage of lipid species but did not measure absolute values of lipid species levels. Although our cohort allowed sex specific analyses, sex differences could not be easily identified due to differences in sociodemographic characteristics between women and men. Because of a relatively small number of HIV-uninfected participants, this study has limited statistical power to detect potential effect modification by HIV infection. Future studies with larger number of both HIV-infected and HIV-uninfected individuals are needed to validate and expand our findings.

In summary, this lipidomic study in two HIV cohorts identified multiple lipid species in plasma associated with development of diabetes, independent of traditional risk factors. Many of these diabetes-associated lipid species were altered in HIV-infected individuals, especially in those using ART. These findings suggest that ART-related plasma lipidomic alterations may precede the development of diabetes among HIV-infected people, which provides new evidence at a lipid species level on the relationships among HIV infection, ART use, dyslipidemia and risk of diabetes.

## Data Availability

Some or all datasets generated during and/or analyzed during the current study are not publicly available but are available from the corresponding author on reasonable request.

## Acknowledgement

The metabolomics profiling was supported by the National Heart, Lung, and Blood Institute (NHLBI) K01HL129892, and other funding sources for this study include NHLBI R01HL140976 and National Institute of Diabetes and Digestive and Kidney Diseases (NIDDK) R01DK119268 to Q.Q., NHLBI R01 HL126543, R01 HL132794, R01HL083760 and R01HL095140 to R.C.K., NHLBI R01HL095129 to W.S.P., NHLBI K01HL137557 to D.B.H., and the New York Regional Center for Diabetes Translation Research (P30 DK111022) from the NIDDK.

Data in this manuscript were collected by the MACS/WIHS Combined Cohort Study (MWCCS). The contents of this publication are solely the responsibility of the authors and do not represent the official views of the National Institutes of Health (NIH). MWCCS (Principal Investigators): Atlanta CRS (Ighovwerha Ofotokun, Anandi Sheth, and Gina Wingood), U01-HL146241-01; Baltimore CRS (Todd Brown and Joseph Margolick), U01-HL146201-01; Bronx CRS (Kathryn Anastos and Anjali Sharma), U01-HL146204-01; Brooklyn CRS (Deborah Gustafson and Tracey Wilson), U01-HL146202-01; Data Analysis and Coordination Center (Gypsyamber D’Souza, Stephen Gange and Elizabeth Golub), U01-HL146193-01; Chicago-Cook County CRS (Mardge Cohen and Audrey French), U01-HL146245-01; Chicago-Northwestern CRS (Steven Wolinsky), U01-HL146240-01; Connie Wofsy Women’s HIV Study, Northern California CRS (Bradley Aouizerat and Phyllis Tien), U01-HL146242-01; Los Angeles CRS (Roger Detels), U01-HL146333-01; Metropolitan Washington CRS (Seble Kassaye and Daniel Merenstein), U01-HL146205-01; Miami CRS (Maria Alcaide, Margaret Fischl, and Deborah Jones), U01-HL146203-01; Pittsburgh CRS (Jeremy Martinson and Charles Rinaldo), U01-HL146208-01; UAB-MS CRS (Mirjam-Colette Kempf and Deborah Konkle-Parker), U01-HL146192-01; UNC CRS (Adaora Adimora), U01-HL146194-01. The MWCCS is funded primarily by the National Heart, Lung, and Blood Institute (NHLBI), with additional co-funding from the Eunice Kennedy Shriver National Institute Of Child Health & Human Development (NICHD), National Human Genome Research Institute (NHGRI), National Institute On Aging (NIA), National Institute Of Dental & Craniofacial Research (NIDCR), National Institute Of Allergy And Infectious Diseases (NIAID), National Institute Of Neurological Disorders And Stroke (NINDS), National Institute Of Mental Health (NIMH), National Institute On Drug Abuse (NIDA), National Institute Of Nursing Research (NINR), National Cancer Institute (NCI), National Institute on Alcohol Abuse and Alcoholism (NIAAA), National Institute on Deafness and Other Communication Disorders (NIDCD), National Institute of Diabetes and Digestive and Kidney Diseases (NIDDK). MWCCS data collection is also supported by UL1-TR000004 (UCSF CTSA), P30-AI-050409 (Atlanta CFAR), P30-AI-050410 (UNC CFAR), and P30-AI-027767 (UAB CFAR).

## References

1. Behrens G, Dejam A, Schmidt H, Balks HJ, Brabant G, Korner T, Stoll M, Schmidt RE. Impaired glucose tolerance, beta cell function and lipid metabolism in HIV patients under treatment with protease inhibitors. Aids. 1999;13(10):F63–70.

2. Carr A, Samaras K, Burton S, Law M, Freund J, Chisholm DJ, Cooper DA. A syndrome of peripheral lipodystrophy, hyperlipidaemia and insulin resistance in patients receiving HIV protease inhibitors. Aids. 1998;12(7):F51–58.

3. Dube MP, Johnson DL, Currier JS, Leedom JM. Protease inhibitor-associated hyperglycaemia. Lancet. 1997;350(9079):713–714.

4. Brown TT, Cole SR, Li X, et al. ANtiretroviral therapy and the prevalence and incidence of diabetes mellitus in the multicenter aids cohort study. Archives of Internal Medicine. 2005;165(10):1179–1184.

5. Tien PC, Schneider MF, Cox C, Karim R, Cohen M, Sharma A, Young M, Glesby MJ. Association of HIV infection with incident diabetes mellitus: impact of using hemoglobin A1C as a criterion for diabetes. Journal of acquired immune deficiency syndromes (1999). 2012;61(3):334–340.

6. Ledergerber B, Furrer H, Rickenbach M, Lehmann R, Elzi L, Hirschel B, Cavassini M, Bernasconi E, Schmid P, Egger M, Weber R, Swiss HIVCS. Factors associated with the incidence of type 2 diabetes mellitus in HIV-infected participants in the Swiss HIV Cohort Study. Clinical infectious diseases : an official publication of the Infectious Diseases Society of America. 2007;45(1):111–119.

7. Brown TT, Glesby MJ. Management of the metabolic effects of HIV and HIV drugs. Nat Rev Endocrinol. 2012;8(1):11–21.

8. Noubissi EC, Katte JC, Sobngwi E. Diabetes and HIV. Curr Diab Rep. 2018;18(11):125.

9. Feeney ER, Mallon PW. HIV and HAART-Associated Dyslipidemia. Open Cardiovasc Med J. 2011;5:49-63.

10. Haffner SM, American Diabetes A. Dyslipidemia management in adults with diabetes. Diabetes Care. 2004;27 Suppl 1:S68-71.

11. Rhee EP, Cheng S, Larson MG, Walford GA, Lewis GD, McCabe E, Yang E, Farrell L, Fox CS, O’Donnell CJ, Carr SA, Vasan RS, Florez JC, Clish CB, Wang TJ, Gerszten RE. Lipid profiling identifies a triacylglycerol signature of insulin resistance and improves diabetes prediction in humans. J Clin Invest. 2011;121(4):1402–1411.

12. Floegel A, Stefan N, Yu Z, Muhlenbruch K, Drogan D, Joost HG, Fritsche A, Haring HU, Hrabe de Angelis M, Peters A, Roden M, Prehn C, Wang-Sattler R, Illig T, Schulze MB, Adamski J, Boeing H, Pischon T. Identification of serum metabolites associated with risk of type 2 diabetes using a targeted metabolomic approach. Diabetes. 2013;62(2):639–648.

13. Drogan D, Dunn WB, Lin W, Buijsse B, Schulze MB, Langenberg C, Brown M, Floegel A, Dietrich S, Rolandsson O, Wedge DC, Goodacre R, Forouhi NG, Sharp SJ, Spranger J, Wareham NJ, Boeing H. Untargeted metabolic profiling identifies altered serum metabolites of type 2 diabetes mellitus in a prospective, nested case control study. Clin Chem. 2015;61(3):487–497.

14. Razquin C, Toledo E, Clish CB, Ruiz-Canela M, Dennis C, Corella D, Papandreou C, Ros E, Estruch R, Guasch-Ferre M, Gomez-Gracia E, Fito M, Yu E, Lapetra J, Wang D, Romaguera D, Liang L, Alonso-Gomez A, Deik A, Bullo M, Serra-Majem L, Salas-Salvado J, Hu FB, Martinez-Gonzalez MA. Plasma Lipidomic Profiling and Risk of Type 2 Diabetes in the PREDIMED Trial. Diabetes Care. 2018;41(12):2617–2624.

15. Merino J, Leong A, Liu CT, Porneala B, Walford GA, von Grotthuss M, Wang TJ, Flannick J, Dupuis J, Levy D, Gerszten RE, Florez JC, Meigs JB. Metabolomics insights into early type 2 diabetes pathogenesis and detection in individuals with normal fasting glucose. Diabetologia. 2018;61(6):1315–1324.

16. Lu J, Lam SM, Wan Q, Shi L, Huo Y, Chen L, Tang X, Li B, Wu X, Peng K, Li M, Wang S, Xu Y, Xu M, Bi Y, Ning G, Shui G, Wang W. High-Coverage Targeted Lipidomics Reveals Novel Serum Lipid Predictors and Lipid Pathway Dysregulation Antecedent to Type 2 Diabetes Onset in Normoglycemic Chinese Adults. Diabetes Care. 2019.

17. Zhao W, Wang X, Deik AA, Hanna DB, Wang T, Haberlen SA, Shah SJ, Lazar JM, Hodis HN, Landay AL, Yu B, Gustafson D, Anastos K, Post WS, Clish CB, Kaplan RC, Qi Q. Elevated Plasma Ceramides Are Associated With Antiretroviral Therapy Use and Progression of Carotid Artery Atherosclerosis in HIV Infection. Circulation. 2019;139(17):2003–2011.

18. Chai JC, Deik AA, Hua S, Wang T, Hanna DB, Xue X, Haberlen SA, Shah SJ, Suh Y, Lazar JM, Gustafson D, Hodis HN, Landay AL, Anastos K, Post WS, Kaplan RC, Clish CB, Qi Q. Association of Lipidomic Profiles With Progression of Carotid Artery Atherosclerosis in HIV Infection. JAMA Cardiol. 2019.

19. Bacon MC, von Wyl V, Alden C, Sharp G, Robison E, Hessol N, Gange S, Barranday Y, Holman S, Weber K, Young MA. The Women’s Interagency HIV Study: an observational cohort brings clinical sciences to the bench. Clin Diagn Lab Immunol. 2005;12(9):1013–1019.

20. Detels R, Jacobson L, Margolick J, Martinez-Maza O, Munoz A, Phair J, Rinaldo C, Wolinsky S. The multicenter AIDS Cohort Study, 1983 to. Public Health. 2012;126(3):196–198.

21. Hanna DB, Post WS, Deal JA, Hodis HN, Jacobson LP, Mack WJ, Anastos K, Gange SJ, Landay AL, Lazar JM, Palella FJ, Tien PC, Witt MD, Xue X, Young MA, Kaplan RC, Kingsley LA. HIV Infection Is Associated With Progression of Subclinical Carotid Atherosclerosis. Clin Infect Dis. 2015;61(4):640–650.

22. Qi Q, Hua S, Clish CB, Scott JM, Hanna DB, Wang T, Haberlen SA, Shah SJ, Glesby MJ, Lazar JM, Burk RD, Hodis HN, Landay AL, Post WS, Anastos K, Kaplan RC. Plasma Tryptophan-Kynurenine Metabolites Are Altered in Human Immunodeficiency Virus Infection and Associated With Progression of Carotid Artery Atherosclerosis. Clin Infect Dis. 2018;67(2):235–242.

23. Paynter NP, Balasubramanian R, Giulianini F, Wang DD, Tinker LF, Gopal S, Deik AA, Bullock K, Pierce KA, Scott J, Martinez-Gonzalez MA, Estruch R, Manson JE, Cook NR, Albert CM, Clish CB, Rexrode KM. Metabolic Predictors of Incident Coronary Heart Disease in Women. Circulation. 2018;137(8):841–853.

24. Kaplan RC, Kingsley LA, Gange SJ, Benning L, Jacobson LP, Lazar J, Anastos K, Tien PC, Sharrett AR, Hodis HN. Low CD4+ T-cell count as a major atherosclerosis risk factor in HIV-infected women and men. AIDS. 2008;22(13):1615–1624.

25. Tien PC, Schneider MF, Cole SR, Levine AM, Cohen M, DeHovitz J, Young M, Justman JE. Antiretroviral therapy exposure and insulin resistance in the Women’s Interagency HIV study. J Acquir Immune Defic Syndr. 2008;49(4):369–376.

26. Slama L, Palella FJ, Jr., Abraham AG, Li X, Vigouroux C, Pialoux G, Kingsley L, Lake JE, Brown TT. Inaccuracy of haemoglobin A1c among HIV-infected men: effects of CD4 cell count, antiretroviral therapies and haematological parameters. J Antimicrob Chemother. 2014;69(12):3360–3367.

27. Brown TT, Li X, Cole SR, Kingsley LA, Palella FJ, Riddler SA, Chmiel JS, Visscher BR, Margolick JB, Dobs AS. Cumulative exposure to nucleoside analogue reverse transcriptase inhibitors is associated with insulin resistance markers in the Multicenter AIDS Cohort Study. AIDS. 2005;19(13):1375–1383.

28. Matthews DR, Hosker JP, Rudenski AS, Naylor BA, Treacher DF, Turner RC. Homeostasis model assessment: insulin resistance and beta-cell function from fasting plasma glucose and insulin concentrations in man. Diabetologia. 1985;28(7):412–419.

29. Hanna DB, Lin J, Post WS, Hodis HN, Xue X, Anastos K, Cohen MH, Gange SJ, Haberlen SA, Heath SL, Lazar JM, Liu C, Mack WJ, Ofotokun I, Palella FJ, Tien PC, Witt MD, Landay AL, Kingsley LA, Tracy RP, Kaplan RC. Association of Macrophage Inflammation Biomarkers With Progression of Subclinical Carotid Artery Atherosclerosis in HIV-Infected Women and Men. J Infect Dis. 2017;215(9):1352–1361.

30. Benjamini Y, Hochberg Y. Controlling the False Discovery Rate - a Practical and Powerful Approach to Multiple Testing. J R Stat Soc B. 1995;57(1):289–300.

31. Braverman NE, Moser AB. Functions of plasmalogen lipids in health and disease. Biochim Biophys Acta. 2012;1822(9):1442–1452.

32. Huynh K, Martins RN, Meikle PJ. Lipidomic Profiles in Diabetes and Dementia. J Alzheimers Dis. 2017;59(2):433–444.

33. Lehmann R, Franken H, Dammeier S, Rosenbaum L, Kantartzis K, Peter A, Zell A, Adam P, Li J, Xu G, Konigsrainer A, Machann J, Schick F, Hrabe de Angelis M, Schwab M, Staiger H, Schleicher E, Gastaldelli A, Fritsche A, Haring HU, Stefan N. Circulating lysophosphatidylcholines are markers of a metabolically benign nonalcoholic fatty liver. Diabetes Care. 2013;36(8):2331–2338.

34. Yea K, Kim J, Yoon JH, Kwon T, Kim JH, Lee BD, Lee HJ, Lee SJ, Kim JI, Lee TG, Baek MC, Park HS, Park KS, Ohba M, Suh PG, Ryu SH. Lysophosphatidylcholine activates adipocyte glucose uptake and lowers blood glucose levels in murine models of diabetes. J Biol Chem. 2009;284(49):33833–33840.

35. Hasegawa H, Lei J, Matsumoto T, Onishi S, Suemori K, Yasukawa M. Lysophosphatidylcholine enhances the suppressive function of human naturally occurring regulatory T cells through TGF-beta production. Biochem Biophys Res Commun. 2011;415(3):526–531.

36. Funai K, Lodhi IJ, Spears LD, Yin L, Song H, Klein S, Semenkovich CF. Skeletal Muscle Phospholipid Metabolism Regulates Insulin Sensitivity and Contractile Function. Diabetes. 2016;65(2):358–370.

37. Nowak C, Salihovic S, Ganna A, Brandmaier S, Tukiainen T, Broeckling CD, Magnusson PK, Prenni JE, Wang-Sattler R, Peters A, Strauch K, Meitinger T, Giedraitis V, Arnlov J, Berne C, Gieger C, Ripatti S, Lind L, Pedersen NL, Sundstrom J, Ingelsson E, Fall T. Effect of Insulin Resistance on Monounsaturated Fatty Acid Levels: A Multi-cohort Non-targeted Metabolomics and Mendelian Randomization Study. PLoS genetics. 2016;12(10):e1006379.

