## Supplementary Tables/Figures for "Plasma lipidomic profiles and risk of diabetes: two prospective cohorts of HIV-infected and HIV-uninfected individuals"

**Supplementary Table 1. Classification of lipid species**

| Lipid classes/sub-classes | Abbreviation | Number of lipid species |
| --- | --- | --- |
| Cholesterol ester | CE | 13 |
| Diacylglycerol | DAG | 15 |
| Lysophosphatidylcholine | LPC | 10 |
| Lysophosphatidylethanolamine | LPE | 8 |
| Monoacylglycerol | MAG | 4 |
| Phosphatidylcholine | PC | 23 |
| Phosphatidylcholine plasmalogen* | PC-P | 15 |
| Phosphatidylethanolamine | PE | 12 |
| Phosphatidylethanolamine plasmalogen* | PE-P | 12 |
| Phosphatidylinositol | PI | 2 |
| Phosphatidylserine | PS | 4 |
| Phosphatidylserine plasmalogen* | PS-P | 3 |
| Sphingolipid † | SL |  |
| Ceramide † | CER | 4 |
| Sphingomyelin † | SM | 11 |
| Sphingosine † | SS | 1 |
| Triacylglycerol | TAG | 74 |
| All lipid species |  | 211 |

* PC-P, PE-P, and PS-P lipid sub-classes were included into PE, PC and PS lipid classes, respectively.

†CER, SM and SS lipid sub-classes were combined into the sphingolipid (SL) lipid class.

CE, cholesteryl ester; CER, ceramide; DAG, diacylglycerol; LPC, lysophosphatidylcholine; LPE, lysophosphatidylethanolamine; MAG, monoacylglycerol; PC, phosphatidylcholine; PC-P, phosphatidylcholine plasmalogen; PE, phosphatidylethanolamine; PE-P, phosphatidylethanolamine plasmalogen; PI, phosphatidylinositol; PS, phosphatidylserine; PS-P, phosphatidylserine plasmalogen; SM, sphingomyelin; SS, sphingosine; TAG, triacylglycerol.

**Supplementary Table 2. Participant characteristics at baseline by cohort and by HIV status**

|  | **Men (MACS)** | | **Women (WIHS)** | |
| --- | --- | --- | --- | --- |
|  | **HIV+** | **HIV-** | **HIV+** | **HIV-** |
| No. of participants | 202 | 101 | 289 | 102 |
| Follow-up (years) | 12.4 (12.0-12.8) | 12.5(12.2-12.7) | 13.1 (8.6-13.5) | 13.0(8.0-13.5) |
| Age (years) | 46 (42-50) | 47 (45-53) | 42 (38-47) | 41.5 (38-46) |
| Race/ethnicity |  |  |  |  |
| White/Other | 115(56.9) | 65(64.4) | 25(8.7) | 5(4.9) |
| Hispanic | 24(11.9) | 10(9.9) | 89(30.8) | 29(28.4) |
| African American | 63(31.2) | 26(25.7) | 175(60.6) | 68(66.7) |
| Education |  |  |  |  |
| Less than High School | 19(9.4) | 5(5) | 118(40.8) | 34(33.3) |
| High School | 27(13.4) | 14(13.9) | 86(29.8) | 31(30.4) |
| College and above | 156(77.2) | 82(81.2) | 85(29.4) | 37(36.3) |
| Current Smoking | 75(37.1) | 24(23.8) | 138(47.8) | 61(21.1)* |
| BMI (Kg/m^2^) | 25.1(22.5-27.9) | 25.8 (23.5-28.3) | 27.2 (24.1-31.5) | 28.8 (24.8-34.1) |
| Fasting glucose | 97 (91-105) | 94.5 (87-101) | 86 (80-92) | 86 (80-93.8) |
| Fasting insulin | 13.1 (10-18.2) | 12.15 (8.8-15.1) | 11 (7-17) | 9 (6-15)* |
| HOMA-IR | 3.25 (2.28-4.66) | 2.7 (2.065-3.8) | 2.3 (1.49-3.48) | 1.9 (1.1-3.16)* |
| Systolic Blood Pressure (mmHg) | 121 (115-129) | 125 (117-131) | 115 (106-124) | 116 (105.3-125.8) |
| Triglyceride | 149.5 (97-220.3) | 112 (76-161) | 112 (77-163) | 85 (68-117)* |
| HDL-cholesterol (mg/dL) | 43.5 (36.1-52) | 49 (41.4-58.1) | 47 (38-57) | 57 (47-68)* |
| Anti-hypertensive medication use | 31(15.3) | 18(17.8) | 50(17.3) | 10(9.8) |
| Lipid lowering medication use | 40(19.8) | 11(10.9) | 12(4.2) | 0(0)* |
| HIV-specific characteristics† |  |  |  |  |
| HIV-infected participants |  | NA |  | NA |
| CD4+ T cell count (cells/mm^3^) | 519 (358-701) | NA | 438 (289-616) | NA |
| Undetectable viral load (<=80 copies/mL) | 134(66.3) | NA | 134(46.4) | NA |
| HIV-1 viral load (copies/mL)‡ | 5615 (1268-32153) | NA | 3700 (680-16000) | NA |
| ART use in past 6 month | 169(83.7) | NA | 213(73.7) | NA |
| PI use | 82(40.6) | NA | 129(44.6) | NA |
| NNRTI use | 94(46.5) | NA | 83(28.7) | NA |
| NRTI use | 166(82.2) | NA | 210(72.7) | NA |

Data are median (IQR) or n (%), assessed at baseline unless otherwise noted. ART indicates antiretroviral therapy; BMI, body mass index; HDL, high-density lipoprotein; HIV, human immunodeficiency virus; IQR, interquartile range; MACS, Multicenter AIDS Cohort Study; WIHS, Women’s Interagency HIV Study.

**P*<0.05 between HIV+ and HIV- participants within each cohort

† HIV-specific characteristics were based on the HIV-infected participants only.

‡Participants with undetectable viral load were excluded.

**Supplementary Table 3 Associations of all 211 lipid species with risk of diabetes**

*(This table is provided as an Excel sheet file due to a large number of lipid species)*

**Supplementary Table 4. Associations of lipid class/subclass summary measures with risk of diabetes**

|  | **Total summary** | | | **Saturated lipid** | | | **Mono-unsaturated lipid** | | | **Poly-unsaturated lipid** | | |
| --- | --- | --- | --- | --- | --- | --- | --- | --- | --- | --- | --- | --- |
|  | # | HR (95% CI) | *P** | # | HR (95% CI) | *P** | # | HR (95% CI) | *P** | # | HR (95% CI) | *P** |
| **Cholesteryl ester (CE)** | | | | | | | | | | | | |
| Model 1 | 13 | 0.98 (0.96,1.00) | 0.21 | 3 | 0.96 (0.89,1.03) | 0.63 | 2 | 0.98 (0.89,1.08) | 0.99 | 8 | 0.96 (0.94,0.99) | 0.02 |
| Model 2 |  | 1.00 (0.98,1.02) | 0.94 |  | 0.99 (0.92,1.07) | 0.87 |  | 1.07 (0.95,1.19) | 0.46 |  | 0.98 (0.95,1.02) | 0.74 |
| **Cerimade (CER)** | | | | | | | | | | | | |
| Model 1 | 4 | 1.01(0.97,1.06) | 0.89 | 3 | 1.02 (0.96,1.08) | 0.73 | 1 | 1.00 (0.85,1.18) | 0.99 | 0 | NA | NA |
| Model 2 |  | 0.98(0.93,1.04) | 0.76 |  | 0.98 (0.92,1.06) | 0.80 |  | 0.92 (0.77,1.10) | 0.46 |  | NA | NA |
| **Diacylglycerol (DAG)** | | | | | | | | | | | | |
| Model 1 | 15 | 1.03(1.02,1.04) | <0.001 | 4 | 1.12 (1.08,1.17) | <0.001 | 3 | 1.17 (1.10,1.23) | <0.001 | 8 | 1.04 (1.02,1.07) | <0.001 |
| Model 2 |  | 1.03 (1.01,1.05) | 0.06 |  | 1.11 (1.05,1.17) | <0.001 |  | 1.17 (1.08,1.26) | <0.001 |  | 1.03 (0.99,1.06) | 0.64 |
| **Lysophosphatidylcholine (LPC)** | | | | | | | | | | | | |
| Model 1 | 10 | 1.01 (0.99,1.03) | 0.82 | 3 | 1.08 (1.02,1.15) | 0.06 | 2 | 1.03 (0.94,1.12) | 0.99 | 5 | 0.99 (0.96,1.03) | 0.86 |
| Model 2 |  | 1.01(0.99,1.03) | 0.76 |  | 1.07 (1.00,1.14) | 0.23 |  | 1.04 (0.95,1.14) | 0.46 |  | 1.00 (0.96,1.05) | 0.98 |
| **Lysophosphatidylethanolamine (LPE)** | | | | | | | | | | | | |
| Model 1 | 8 | 1.00(0.97,1.02) | 0.90 | 4 | 1.03 (0.98,1.08) | 0.63 | 1 | 0.99 (0.85,1.16) | 0.99 | 3 | 0.94 (0.88,1.00) | 0.15 |
| Model 2 |  | 1.00(0.97,1.03) | 0.96 |  | 1.03 (0.98,1.09) | 0.56 |  | 1.05 (0.88,1.26) | 0.60 |  | 0.92 (0.85,0.99) | 0.31 |
| **Monoacylglycerol (MAG)** | | | | | | | | | | | | |
| Model 1 | 4 | 1.00 (0.95,1.06) | 0.92 | 1 | 1.00 (0.86,1.17) | 0.96 | 3 | 1.00 (0.94,1.08) | 0.99 | 0 | NA | NA |
| Model 2 |  | 0.96(0.91,1.02) | 0.76 |  | 0.92 (0.78,1.08) | 0.56 |  | 0.95 (0.88,1.02) | 0.41 |  | NA | NA |
| **Phosphatidylcholine (PC)** | | | | | | | | | | | | |
| Model 1 | 23 | 1.01(1.00,1.01) | 0.74 | 4 | 1.04 (0.99,1.09) | 0.34 | 4 | 1.04 (1.00,1.09) | 0.16 | 15 | 1.00 (0.99,1.02) | 0.78 |
| Model 2 |  | 1.00(0.99,1.02) | 0.76 |  | 1.04 (0.99,1.10) | 0.43 |  | 1.04 (0.99,1.10) | 0.29 |  | 1.00 (0.98,1.02) | 0.98 |
| **Phosphatidylethanolamine plasmalogen (PC-P)** | | | | | | | | | | | | |
| Model 1 | 15 | 0.99 (0.97,1.00) | 0.28 | 0 | NA | NA | 3 | 0.94 (0.88,0.99) | 0.11 | 12 | 0.99 (0.97,1.00) | 0.25 |
| Model 2 |  | 0.99 (0.98,1.01) | 0.76 |  | NA | NA |  | 0.97 (0.91,1.04) | 0.46 |  | 0.99 (0.97,1.01) | 0.98 |
| **Phosphatidylethanolamine (PE)** | | | | | | | | | | | | |
| Model 1 | 12 | 1.01(1.00,1.03) | 0.37 | 2 | 1.03 (0.95,1.12) | 0.73 | 1 | 1.26 (1.08,1.48) | 0.02 | 9 | 1.02 (0.99,1.04) | 0.32 |
| Model 2 |  | 1.01 (0.99,1.04) | 0.76 |  | 1.03 (0.94,1.12) | 0.80 |  | 1.28 (1.05,1.57) | 0.07 |  | 1.01 (0.99,1.04) | 0.74 |
| **Phosphatidylethanolamine plasmalogen (PE-P)** | | | | | | | | | | | | |
| Model 1 | 12 | 0.99(0.98,1.01) | 0.82 | 0 | NA | NA | 0 | NA | NA | 12 | 0.99 (0.98,1.01) | 0.70 |
| Model 2 |  | 1.00(0.98,1.02) | 0.96 |  | NA | NA |  | NA | NA |  | 1.00 (0.98,1.02) | 0.98 |
| **Phosphatidylinositol (PI)** | | | | | | | | | | | | |
| Model 1 | 2 | 0.98 (0.89,1.08) | 0.89 | 1 | 0.97 (0.83,1.14) | 0.81 | 1 | 0.98 (0.83,1.15) | 0.99 | 0 | NA | NA |
| Model 2 |  | 0.96 (0.86,1.08) | 0.76 |  | 0.98 (0.82,1.17) | 0.87 |  | 0.92 (0.76,1.11) | 0.46 |  | NA | NA |
| **Phosphatidylserine (PS)** | | | | | | | | | | | | |
| Model 1 | 4 | 1.00(0.95,1.06) | 0.92 | 1 | 1.04 (0.89,1.22) | 0.78 | 0 | NA | NA | 3 | 1.00 (0.93,1.06) | 0.94 |
| Model 2 |  | 1.00(0.94,1.06) | 0.96 |  | 1.05 (0.88,1.25) | 0.80 |  | NA | NA |  | 0.99 (0.92,1.07) | 0.98 |
| **Phosphatidylserine plasmalogen (PS-P)** | | | | | | | | | | | | |
| Model 1 | 3 | 1.01 (0.95,1.08) | 0.89 | 0 | NA | NA | 1 | 1.02 (0.87,1.19) | 0.99 | 2 | 1.02 (0.93,1.12) | 0.78 |
| Model 2 |  | 1.02 (0.95,1.10) | 0.76 |  | NA | NA |  | 1.11 (0.93,1.33) | 0.46 |  | 1.01 (0.90,1.12) | 0.98 |
| **Sphingomyelin (SM)** | | | | | | | | | | | | |
| Model 1 | 11 | 0.99 (0.98,1.01) | 0.82 | 6 | 0.99 (0.96,1.02) | 0.73 | 4 | 0.98 (0.94,1.03) | 0.83 | 1 | 0.94 (0.80,1.09) | 0.68 |
| Model 2 |  | 0.99(0.97,1.01) | 0.76 |  | 0.99 (0.96,1.03) | 0.80 |  | 0.98 (0.93,1.03) | 0.46 |  | 0.93 (0.79,1.09) | 0.74 |
| **Sphingosine (SS)** | | | | | | | | | | | | |
| Model 1 | 1 | 0.97(0.83,1.13) | 0.89 | 1 | 0.97 (0.83,1.13) | 0.78 | 0 | NA | NA | 0 | NA | NA |
| Model 2 |  | 0.92(0.78,1.08) | 0.76 |  | 0.92 (0.78,1.08) | 0.56 |  | NA | NA |  | NA | NA |
| **Triacylglycerol (TAG)** | | | | | | | | | | | | |
| Model 1 | 74 | 1.01(1.00,1.01) | <0.001 | 10 | 1.04 (1.02,1.06) | 0.00 | 11 | 1.04 (1.02,1.05) | <0.001 | 53 | 1.01 (1.00,1.01) | 0.01 |
| Model 2 |  | 1.01(1.00,1.01) | 0.17 |  | 1.03 (1.01,1.05) | 0.02 |  | 1.03 (1.01,1.05) | 0.02 |  | 1.00 (1.00,1.01) | 0.64 |

Data are risk ratios (HRs) and 95% confidence intervals (CIs) of incident diabetes per unit increase of lipid class/subclass summary scores, stratified by sex and adjusted for age, race/ethnicity, education, study site, current smoking, HIV serostatus and treatment status (HIV-, HIV+ ART user, HIV+ ART non-user) and CD4 cell counts (Model 1); and further adjusted for systolic blood pressure, HDL-cholesterol, triglycerides, BMI, anti-hypertensive medication use and lipid lowering medication use (Model 2). The summary scores were calculated as the summation of inverse-normalized values of all lipid species, saturated lipid species, mono-unsaturated lipid species, or poly-unsaturation lipid species within each class/subclass. #indicates number of lipid species included in each category.

^*^P-values are FDR adjusted across 16 subclasses

**Supplementary Table 5. Separate and joint analyses of top and secondary lipids in each lipid class/subclass**

| Lipid class | Lipid species | | r | Separate analysis | | Joint analysis | |
| --- | --- | --- | --- | --- | --- | --- | --- |
|  |  |  |  | HR (%95 CI) | *P*-values | HR (%95 CI) | *P*-values |
| LPC | Top LPC | LPC (18:2) | 0.349 | 0.80 (0.69,0.94) | 0.007 | 0.72 (0.61,0.86) | <0.001 |
|  | Secondary LPC | LPC (14:0) |  | 1.24 (1.05,1.47) | 0.010 | 1.40 (1.17,1.67) | <0.001 |
| PC | Top PC | PC (38:3) | 0.231 | 1.25 (1.07,1.46) | 0.005 | 1.33 (1.13,1.56) | <0.001 |
|  | Secondary PC | PC (36:4) |  | 0.84 (0.72,0.98) | 0.027 | 0.78 (0.67,0.92) | 0.003 |
| PE | Top PE | PE (36:1) | 0.321 | 1.26 (1.07,1.47) | 0.005 | 1.47 (1.23,1.75) | <0.001 |
|  | Secondary PE | PE (38:2) |  | 0.81 (0.69,0.96) | 0.012 | 0.70 (0.59,0.83) | <0.001 |

Hazards Ratios (HRs) and 95% confidence intervals (CIs) of risk diabetes per SD increment of lipid species were estimated, stratified by sex and adjusted for age, race/ethnicity, education and current smoking. Pearson correlation coefficients (r) between top and secondary lipid species in the same lipid class/subclass were estimated.

**Supplementary Table 6. Associations of 11 featured lipid species with risk of diabetes in men and women separately as well as combined using a meta-analysis approach**

| **Lipid species** | **Men (N=303)** | |  | **Women (N=391)** | |  | *P* for interaction |  | **Combined by Meta-analysis** | | |
| --- | --- | --- | --- | --- | --- | --- | --- | --- | --- | --- | --- |
|  | HR (95%CI) | *P*-value |  | HR (95%CI) | *P*-value |  |  |  | HR (95%CI) | *P*-value | *P* for heterogeneity |
| CE (22:4) |  |  |  |  |  |  |  |  |  |  |  |
| Model 1 | 0.58 (0.44,0.76) | <0.001 |  | 0.67 (0.55,0.83) | <0.001 |  | 0.60 |  | 0.64 (0.54,0.75) | <0.001 | 0.38 |
| Model 2 | 0.74 (0.53,1.03) | 0.07 |  | 0.72 (0.56,0.94) | 0.01 |  | 0.61 |  | 0.73 (0.60,0.90) | 0.003 | 0.94 |
| DAG (32:0) |  |  |  |  |  |  |  |  |  |  |  |
| Model 1 | 1.72 (1.32,2.25) | <0.001 |  | 1.57 (1.27,1.94) | <0.001 |  | 0.63 |  | 1.63 (1.38,1.92) | <0.001 | 0.58 |
| Model 2 | 1.30 (0.92,1.83) | 0.12 |  | 1.68 (1.26,2.23) | <0.001 |  | 0.90 |  | 1.52 (1.22, 1.89) | <0.001 | 0.27 |
| LPC (18:2)* |  |  |  |  |  |  |  |  |  |  |  |
| Model 1 | 0.63 (0.47,0.84) | <0.001 |  | 0.78 (0.63,0.98) | 0.03 |  | 0.23 |  | 0.73 (0.61, 0.87) | <0.001 | 0.23 |
| Model 2 | 0.83 (0.60,1.14) | 0.26 |  | 0.86 (0.67,1.11) | 0.27 |  | 0.44 |  | 0.85 (0.70,1.04) | 0.12 | 0.84 |
| LPC (14:0)* |  |  |  |  |  |  |  |  |  |  |  |
| Model 1 | 1.20 (0.91,1.60) | 0.18 |  | 1.56 (1.22,1.98) | <0.001 |  | 0.22 |  | 1.40 (1.17,1.68) | <0.001 | 0.18 |
| Model 2 | 0.90 (0.66,1.23) | 0.51 |  | 1.46 (1.14,1.88) | <0.001 |  | 0.10 |  | 1.21 (1.00,1.47) | 0.05 | 0.02 |
| PC (P-34:3) |  |  |  |  |  |  |  |  |  |  |  |
| Model 1 | 0.58 (0.45,0.76) | <0.001 |  | 0.85 (0.69,1.05) | 0.15 |  | 0.05 |  | 0.74 (0.63, 0.88) | <0.001 | 0.03 |
| Model 2 | 0.70 (0.51,0.96) | 0.02 |  | 0.88 (0.68,1.15) | 0.37 |  | 0.07 |  | 0.81 (0.66,0.99) | 0.04 | 0.27 |
| PC (38:3)* |  |  |  |  |  |  |  |  |  |  |  |
| Model 1 | 1.53 (1.14,2.06) | <0.001 |  | 1.33 (1.05,1.69) | 0.01 |  | 0.58 |  | 0.75 (0.63,0.89) | 0.001 | 0.32 |
| Model 2 | 1.17 (0.84,1.64) | 0.34 |  | 1.20 (0.90,1.59) | 0.19 |  | 0.62 |  | 0.83 (0.68,1.00) | 0.06 | 0.63 |
| PC (36:4) |  |  |  |  |  |  |  |  |  |  |  |
| Model 1 | 0.67 (0.50,0.89) | <0.001 |  | 0.80 (0.64,0.99) | 0.04 |  | 0.38 |  | 1.41 (1.17,1.70) | <0.001 | 0.47 |
| Model 2 | 0.77 (0.56,1.07) | 0.12 |  | 0.85 (0.66,1.09) | 0.21 |  | 0.30 |  | 1.19 (0.96,1.48) | 0.11 | 0.91 |
| PE (36:1)* |  |  |  |  |  |  |  |  |  |  |  |
| Model 1 | 1.47 (1.11,1.96) | <0.001 |  | 1.47 (1.15,1.88) | <0.001 |  | 0.83 |  | 1.48 (1.23,1.77) | <0.001 | 0.99 |
| Model 2 | 1.25 (0.90,1.74) | 0.17 |  | 1.40 (1.04,1.87) | 0.02 |  | 0.63 |  | 1.34 (1.07,1.66) | 0.01 | 0.62 |
| PE (38:2) |  |  |  |  |  |  |  |  |  |  |  |
| Model 1 | 0.71 (0.53,0.96) | 0.03 |  | 0.68 (0.54,0.86) | <0.001 |  | 0.96 |  | 0.70 (0.58,0.84) | <0.001 | 0.82 |
| Model 2 | 0.71 (0.50,1.00) | 0.05 |  | 0.77 (0.60,0.98) | 0.04 |  | 0.42 |  | 0.75 (0.62,0.92) | 0.01 | 0.71 |
| PE (P-38:3) |  |  |  |  |  |  |  |  |  |  |  |
| Model 1 | 0.59 (0.44,0.78) | <0.001 |  | 0.93 (0.75,1.15) | 0.53 |  | 0.02 |  | 0.79 (0.67,0.94) | 0.01 | 0.01 |
| Model 2 | 0.72 (0.54,0.97) | 0.03 |  | 1.02 (0.81,1.28) | 0.83 |  | 0.01 |  | 0.90 (0.75,1.08) | 0.25 | 0.07 |
| TAG (50:1) |  |  |  |  |  |  |  |  |  |  |  |
| Model 1 | 1.75 (1.35,2.28) | <0.001 |  | 1.46 (1.18,1.80) | <0.001 |  | 0.34 |  | 1.57 (1.33,1.85) | <0.001 | 0.28 |
| Model 2 | 1.35 (0.96,1.90) | 0.07 |  | 1.49 (1.12,1.97) | <0.001 |  | 0.49 |  | 1.44 (1.15,1.78) | <0.001 | 0.68 |

Data are risk ratios (HRs) and 95% confidence intervals (CIs) of incident diabetes per SD increment of lipid species, adjusted for age, race/ethnicity, education, study site, current smoking, HIV serostatus and treatment status (HIV-, HIV+ ART user, HIV+ ART non-user) and CD4 cell counts (Model 1); and further adjusted for systolic blood pressure, HDL-cholesterol, triglycerides, BMI, anti-hypertensive medication use and lipid lowering medication use (Model 2). HRs estimated in men and women separately were combined by fixed-effect meta-analysis

*Top and Secondary lipid species within the same lipid classes/subclasses were included in the Cox models simultaneously to estimate RRs (95% CI) and *P*-values

CE, cholesteryl ester; DAG, diacylglycerol; LPC, lysophosphatidylcholine; PC, phosphatidylcholine; PC(P), phosphatidylcholine plasmalogen; PE, phosphatidylethanolamine; PE (P), phosphatidylethanolamine plasmalogen; TAG, triacylglycerol.

**Supplementary Table 7. Comparison in diabetes-associated lipid species between HIV-infected and HIV-uninfected participants**

|  | CE (22:4) | DAG (32:0) | LPC (18:2) | LPC (14:0) | PC (P-34:3) | PC (38:3) | PC (36:4) | PE (36:1) | PE (38:2) | PE (P-38:3) | TAG (50:1) |
| --- | --- | --- | --- | --- | --- | --- | --- | --- | --- | --- | --- |
| Beta (SE) | 0.316 (0.082) | 0.296  (-0.082) | -0.175  (-0.082) | 0.123  (-0.084) | -0.527  (-0.081) | 0.684  (-0.079) | -0.227  (-0.082) | 0.342  (-0.081) | 0.016  (-0.082) | -0.507  (-0.081) | 0.302  (-0.083) |
| *P*-value | <0.001 | <0.001 | 0.034 | 0.143 | <0.001 | <0.001 | 0.006 | <0.001 | 0.844 | <0.001 | <0.001 |

Data are differences (beta [SE]) in lipid species (inverse normalized) between HIV-infected participants and HIV-uninfected participants, adjusted for age and sex.

CE, cholesteryl ester; DAG, diacylglycerol; LPC, lysophosphatidylcholine; PC, phosphatidylcholine; PC(P), phosphatidylcholine plasmalogen; PE, phosphatidylethanolamine; PE (P), phosphatidylethanolamine plasmalogen; TAG, triacylglycerol.

**Supplementary Figure 1. Manhattan plot for the associations of 211 lipid species with risk of diabetes.**

Individual lipid species are depicted by filled circles and arranged by lipid class in 11 panels according to the number of total carbon atoms (x axes) and –log10 P-values (y axes). Dash line represents cut-off of FDR<0.05. Diamond shapes with lipid names are the top lipid species associated with risk of incident diabetes in each of the lipid classes. Circle color indicates Spearman correlation coefficients between the top lipid species and other lipid species within each lipid class.

CE, cholesteryl ester; CER, ceramide; DAG, diacylglycerol; LPC, lysophosphatidylcholine; LPE, lysophosphatidylethanolamine; MAG, monoacylglycerol; PC, phosphatidylcholine; PE, phosphatidylethanolamine; PI, phosphatidylinositol; PS, phosphatidylserine; SL, sphingolipid, TAG, triacylglycerol.

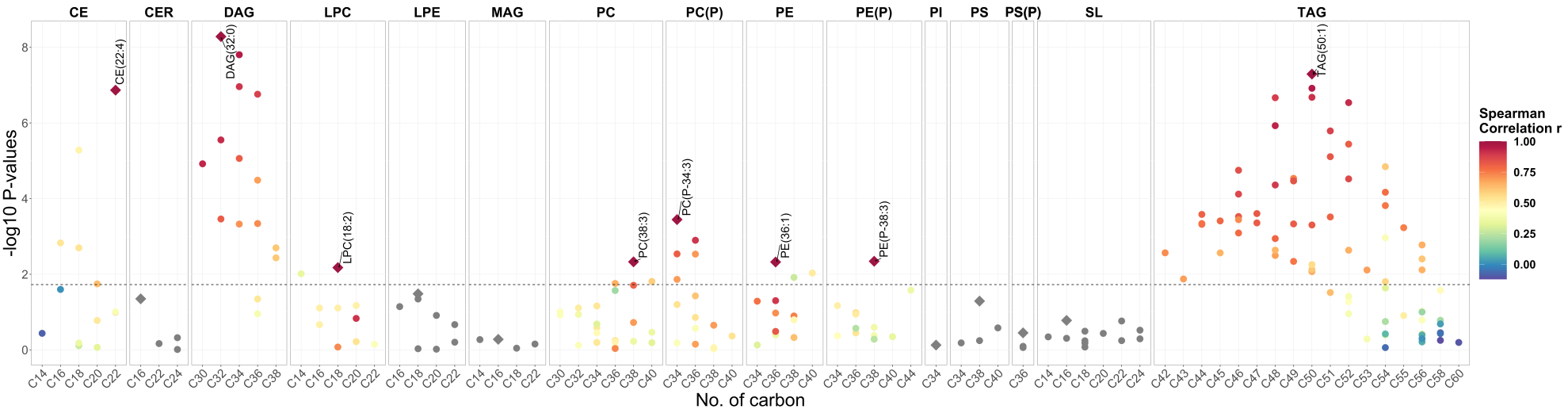

**Supplementary Figure 2. The flow chart for the selection of top and secondary lipids in each lipid class/subclass.**

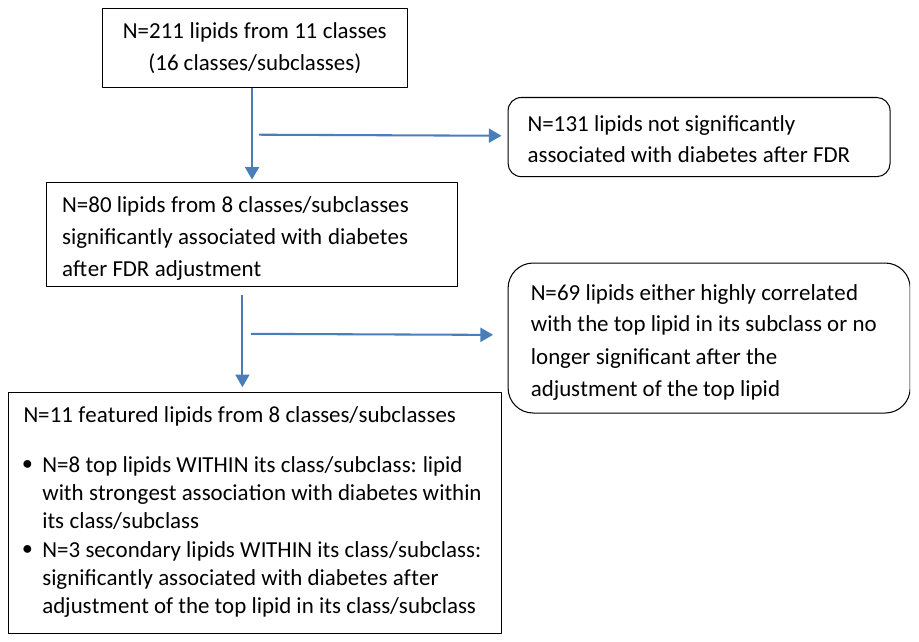

**Supplementary Figure 3. Partial Spearman correlation of lipid species with diabetes and HIV related factors in HIV-infected and HIV-uninfected participants separately**

Data are partial Spearman correlation coefficients (r) of 11 diabetes-associated lipid species levels with age, BMI, fasting glucose, fasting insulin, HOMR-IR, HDL-cholesterol and triglycerides (excluding participants taking lipid-lowering medication) and systolic blood pressure (excluding participants taking anti-hypertensive medication) by HIV serostatus group. * *P*-value < 0.05.

BMI, body mass index; CE, cholesteryl ester; DAG, diacylglycerol; LPC, lysophosphatidylcholine; PC, phosphatidylcholine; PC(P), phosphatidylcholine plasmalogen; PE, phosphatidylethanolamine; PE (P), phosphatidylethanolamine plasmalogen; TAG, triacylglycerol.

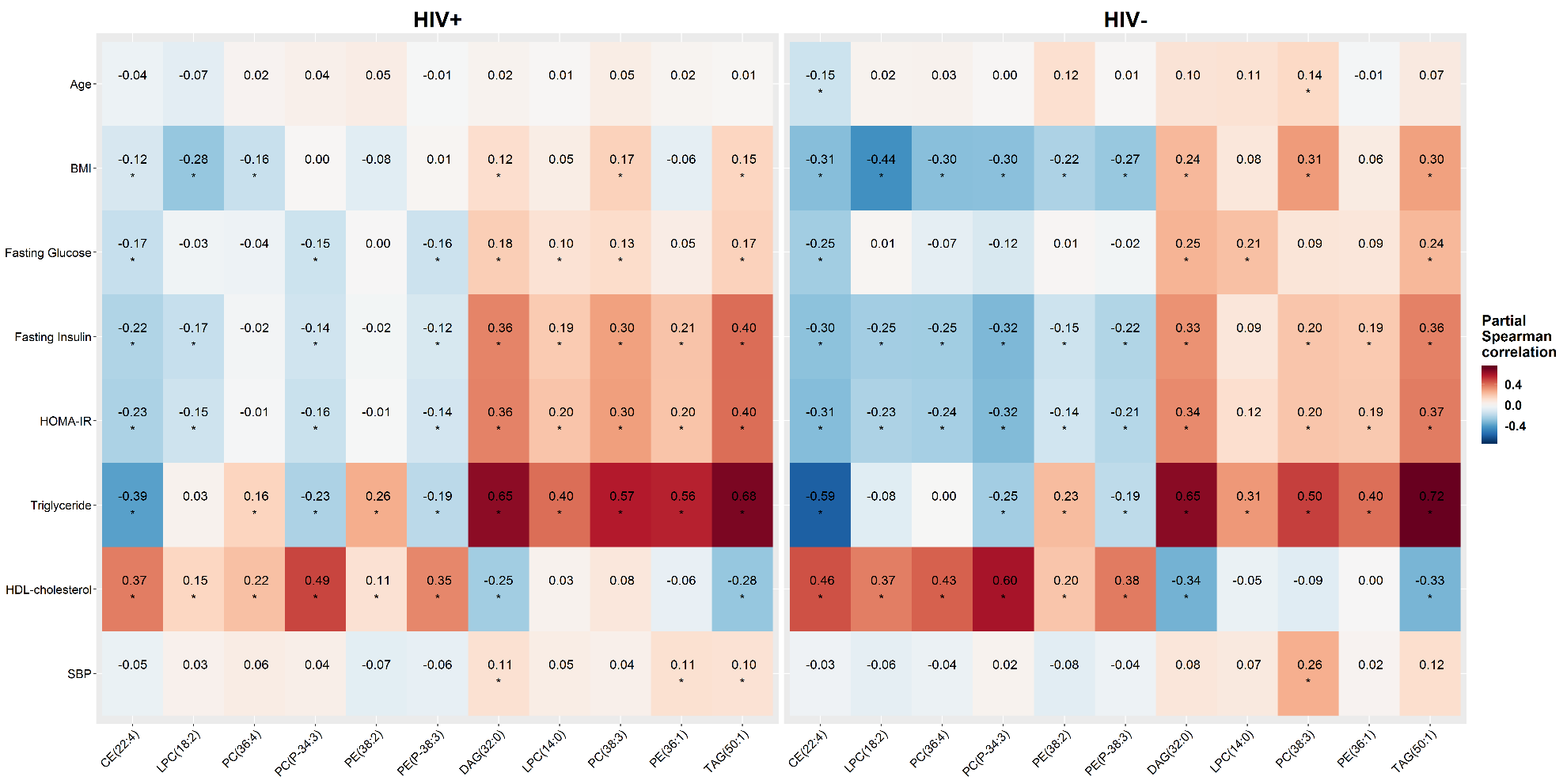

**Supplementary Figure 4 Plasma levels of diabetes associated lipid species according to HIV infection and classes of ART use.**

Data are inverse normal transformed levels of diabetes-associated lipid species in HIV-uninfected participants (group 0, blue), HIV-infected participants without ART subclass use (group 1, green), and HIV-infected participants with ART subclass use (group 2, red). A, B, and C represent PI, NRTI, and NNRTI subclass, respectively.

ART, antiretroviral therapy; CE, cholesteryl ester; CER, ceramide; DAG, diacylglycerol; LPC, lysophosphatidylcholine; LPE, lysophosphatidylethanolamine; MAG, monoacylglycerol; PC, phosphatidylcholine; PE, phosphatidylethanolamine; PI, phosphatidylinositol; PS, phosphatidylserine; TAG, triacylglycerol.

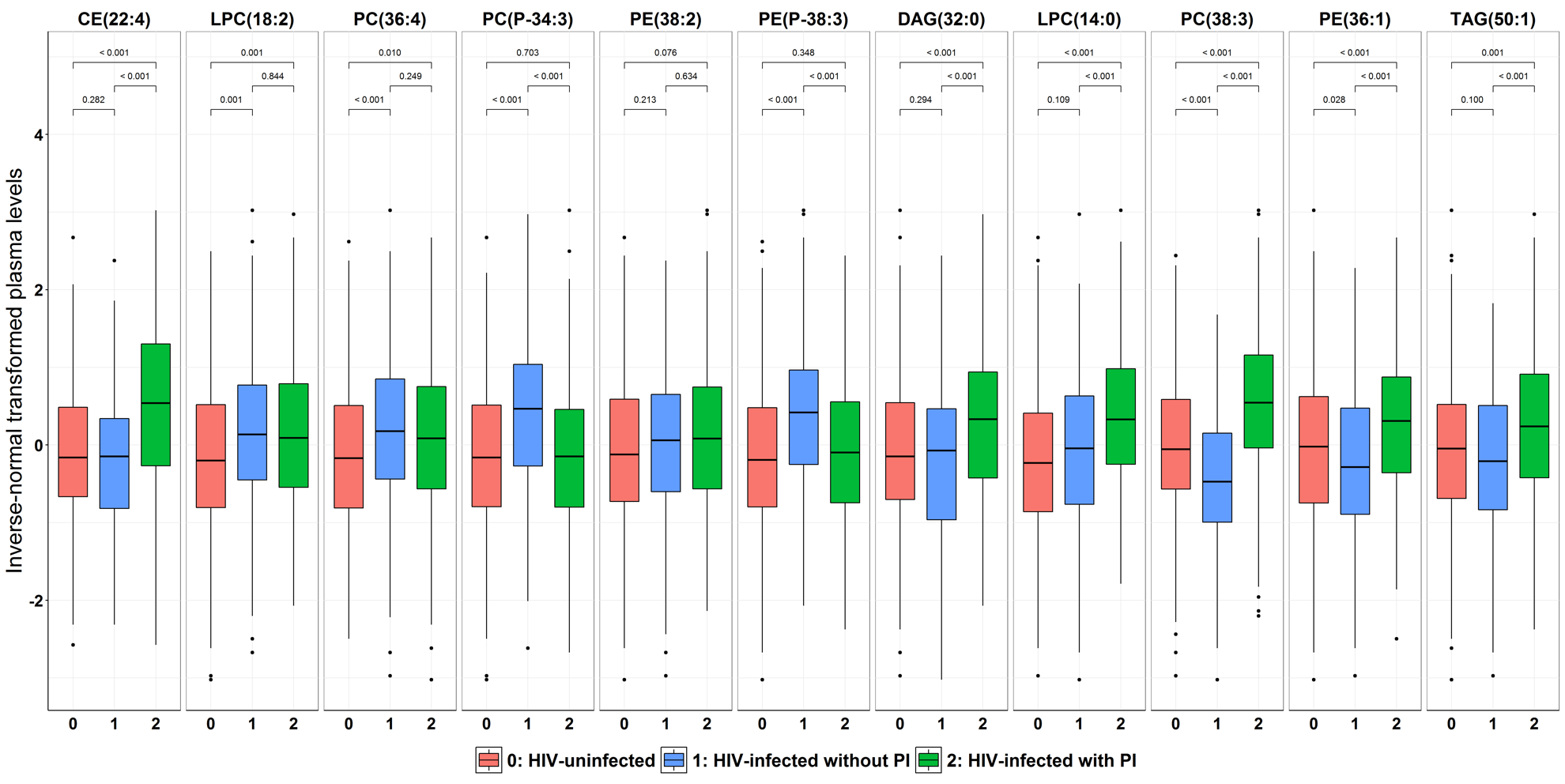

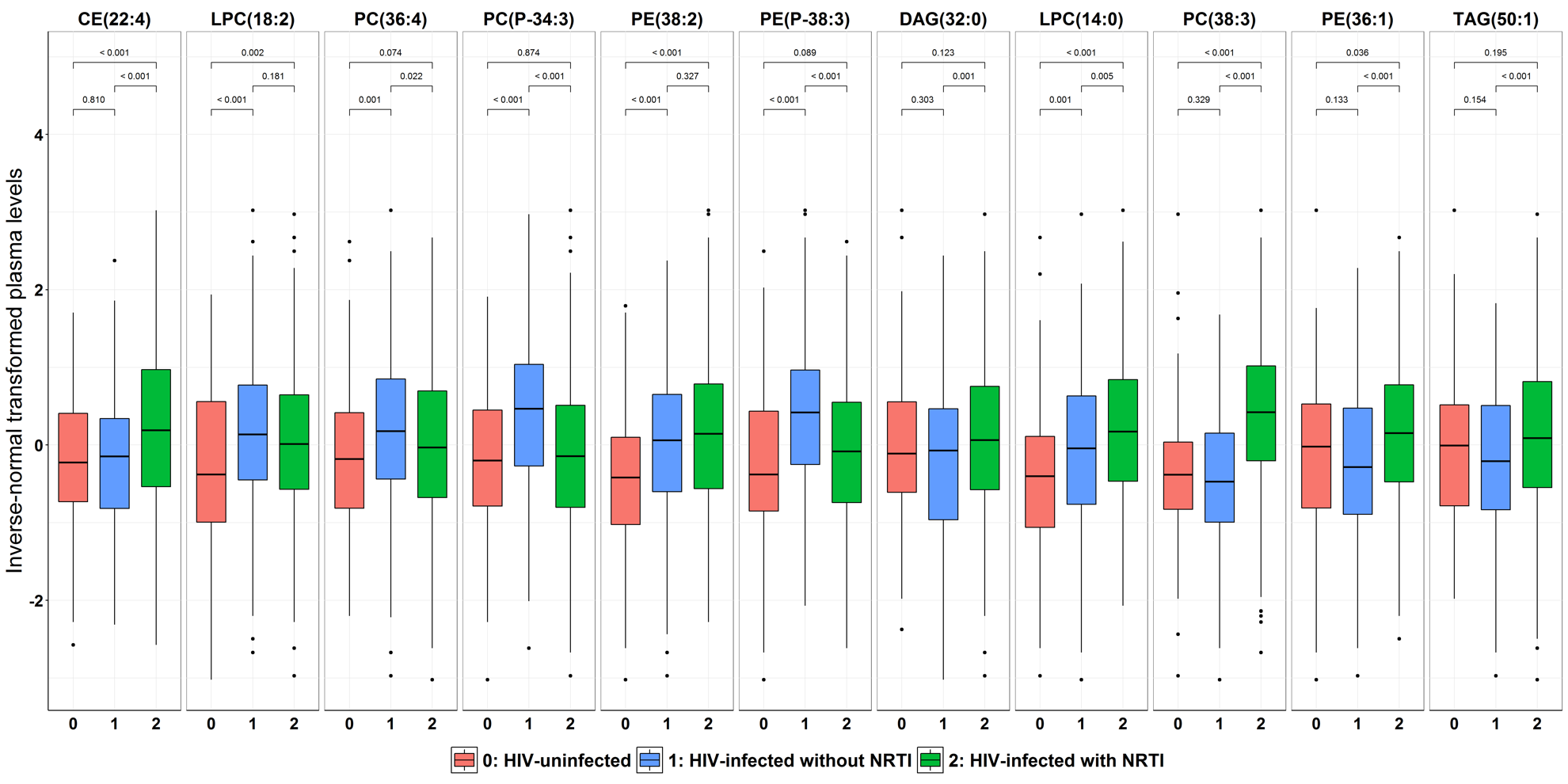

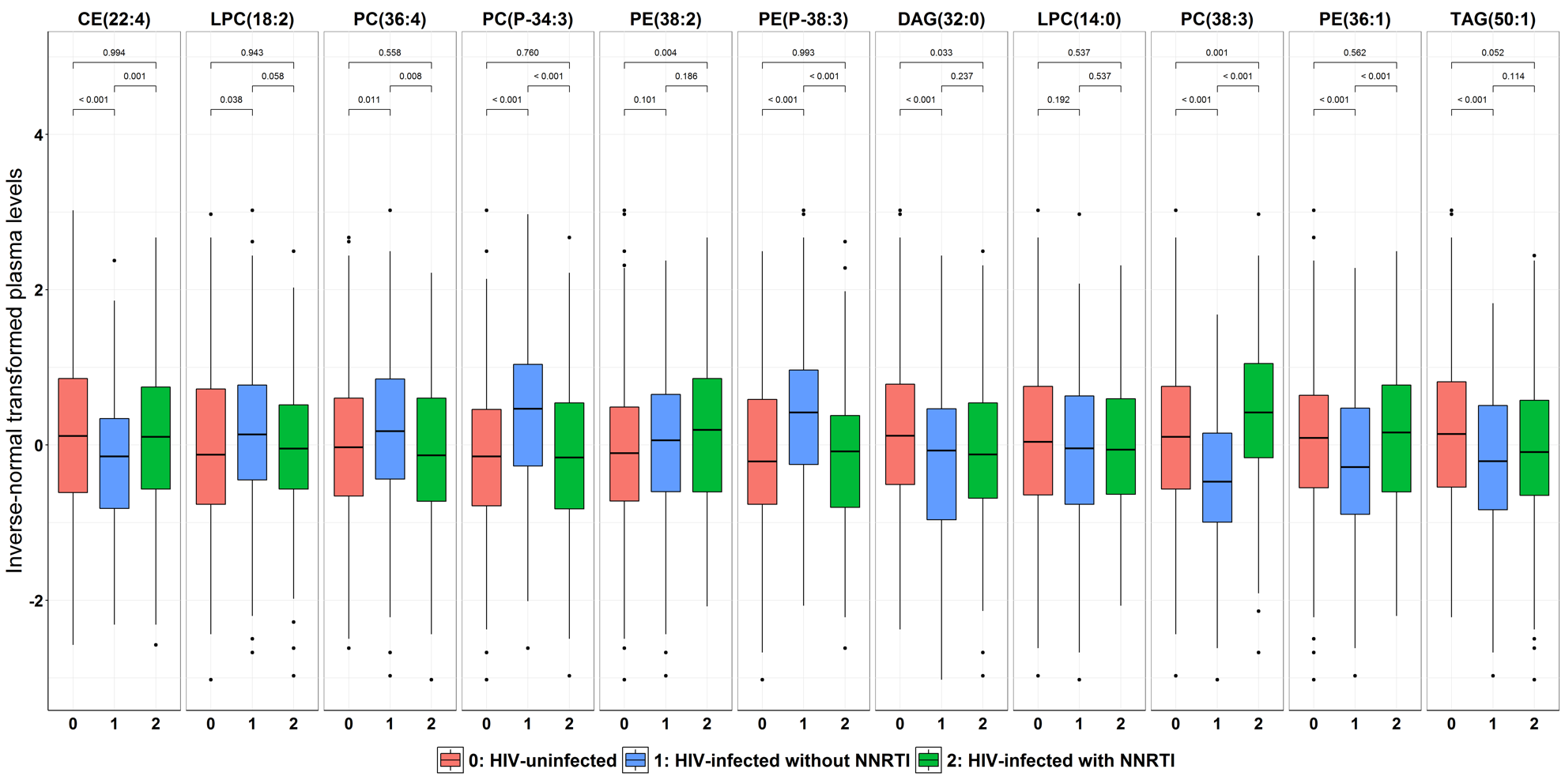
